# Large-Scale Psychiatric Concept Extraction from Electronic Health Records: A Comparative Study of Encoder-Based Language Models

**DOI:** 10.64898/2026.08.20.26360921

**Authors:** Xiaodong Xue, Clara Frydman-Gani, Alejandro Arias, Maria Perez Vallejo, John Daniel Londoño Martínez, Johanna Valencia-Echeverry, Mauricio Castaño, Nelson B. Freimer, Carlos Lopez-Jaramillo, Loes M. Olde Loohuis

**Author notes:** To whom correspondence should be addressed: Dr. Loes Olde Loohuis, Psychiatry & Biobehavioral Sciences at the Center for Neurobehavioral Genetics Gonda Center, 695 Charles E. Young Drive South Los Angeles, CA 90095. Equal contribution (Xiaodong Xue, Clara Frydman-Gani).

## Abstract

**Background:** Free-text notes in electronic health records (EHRs) contain fine-grained psychiatric information that is essential for psychiatric research and clinical care, and often absent or under-recorded in structured codes alone. Clinical natural language processing (cNLP) can support extraction of this information from EHR notes, yet Spanish-language cNLP remains under-developed. Moreover, broad evaluations comparing multiple encoder-based language models across extensive, fine-grained psychiatric concept sets remain scarce, and it remains unclear how these models compare with traditional NLP (tNLP) systems and much larger generative large language models (LLMs). In addition, cross-site performance of fine-tuned models is rarely tested, and limited annotated training data remains a major challenge, especially for rare symptoms.

**Objectives:** We aimed to advance scalable, global psychiatric cNLP by fine-tuning multiple encoder-based models with differing architectures and pre-training strategies for detecting fine-grained psychiatric concepts in Spanish EHRs. We further evaluated the impact of augmenting the fine-tuning data with precision-weighted weak labels for less-frequent concepts, and compared the performance of the encoder-based models to that of tNLP and a fine-tuned generative LLM trained on the same data. Finally, we evaluated model cross-site generalizability on an external EHR dataset.

**Methods:** Three encoder-based models (BETO, XLM-RoBERTa-large, and bsc-bio-ehr-es) were fine-tuned on 1,642 clinician-annotated EHR documents from Colombia to detect 110 psychiatric concepts in Spanish text. To address the limited annotated examples available for less-frequent concepts, 12,000 additional documents were weakly-labeled for less-frequent concepts using tNLP, and incorporated into the fine-tuning data with labels weighted by pattern precision. Models were compared with tNLP and a generative LLM, and evaluated on an external EHR dataset from another psychiatric hospital in Colombia.

**Results:** Encoder model performance varied substantially, with macro-F1 ranging from 0.64 to 0.81. BETO achieved the highest macro-F1 (0.81; median F1=0.88 [IQR=0.77-0.96]). Adding precision-weighted weak labels for less-frequent concepts improved BETO’s overall macro-F1 to 0.83 and increased mean F1 for the 55 augmented concepts from 0.82 to 0.86. Under matched fine-tuning conditions, fine-tuned BETO and the tNLP method were equivalent in F1, whereas the LLM significantly outperformed BETO in F1. After weak-label augmentation, BETO significantly outperformed tNLP in F1 (*P_FDR_*<.001) and narrowed the performance gap with the LLM, although equivalence was not established. Lastly, fine-tuned BETO maintained reasonably strong performance on data from an external hospital not used for model fine-tuning (out-of-domain macro-F1=0.78).

**Conclusions:** General-purpose pre-trained encoders had strong performance for psychiatric concept extraction from Spanish EHRs. Weak-label augmentation improved BETO’s performance and strengthened results relative to a tNLP baseline, while reducing, but not eliminating, the performance gap with a much larger fine-tuned generative LLM. These findings highlight the utility of these relatively lightweight models for scalable, accurate and reproducible detection of psychiatric concepts in Spanish-language EHRs.

## Introduction

The accurate extraction of clinical phenotypes from electronic health records (EHRs) is essential for a wide range of applications in clinical studies and precision medicine, including cohort assembly, medical history summarization, prediction of clinical outcomes, and longitudinal analyses [1–4]. In psychiatry, where reliable biomarkers remain limited, most symptom-level information documented in EHRs - including features like depressed mood, anhedonia, hallucinations, and suicidal ideation - is embedded in free-text notes recorded by clinicians[5]. This information can be used to improve psychiatric cases and control definitions for clinical and genetic research[1], predict psychiatric conditions from history of present illness[3], and enhance the prediction of mental health crises and hospitalization[2,4]. Automated concept extraction enables these analyses to extend from small, manually-reviewed cohorts to larger cohort- and population-level studies, supporting patient stratification, tracking of features over time, and large-scale epidemiologic and genetic studies. Because these features are often absent or under-recorded in structured EHR data[6–11], the extraction of fine-grained psychiatric concepts from clinical text is particularly valuable.

Given the unstructured nature of EHR notes, the extensive and scalable extraction of information from these notes remains challenging[12–14]. Successful extraction methods must contend with variable documentation practices, heterogeneous language, and clinical shorthand. Many clinical NLP (cNLP) methods - that is, computational methods for analyzing and extracting information from clinical text - have been developed to address these challenges, ranging from pattern-based systems to advanced language models based on the Transformer[15] architecture. Traditional NLP (tNLP) approaches, defined here as methods that identify relevant expressions using predefined patterns and lexicons, have enabled fast, scalable and interpretable detection of psychiatric concepts from EHR notes[16–19]. For example, the UK-based CRIS-CODE project developed applications for the extraction of over 50 psychiatric symptoms from English clinical records[16]; other systems identified suicidal ideation and suicide attempts[17], classified mood state and treatment-resistant depression[18], or incorporated information extracted from notes into post-discharge suicide-risk predictions[19]. However, most of these studies have generally focused on a narrow set of symptoms and clinical outcomes, and lack cross-institutional generalizability evaluations. Moreover, tNLP methods can be limited in their ability to generalize across linguistic variability and recognize contextual nuance.

With the introduction of the Transformer architecture, encoder-only masked language models - such as BERT[20] and RoBERTa[21] - have been utilized for this purpose as well. These models utilize the Transformer’s encoder to process the full input sequence bidirectionally to learn contextual representations, and are pre-trained on massive corpora of text - for example, BERT was pre-trained on BookCorpus and English Wikipedia, whereas RoBERTa was further pre-trained CC-News, OpenWebText, and web-based stories[20,21] - to gain linguistic and contextual knowledge. These encoder-only models can then be fine-tuned for specific tasks, including classification and named entity recognition. While more resource-intensive than tNLP methods (i.e., requiring greater memory and computational capacity for fine-tuning and inference), Transformer-based models offer advantages through their context-sensitive token embeddings that better capture nuanced semantic expressions, and adaptability across diverse clinical applications[22–25]. Encoders have been applied to various psychiatric tasks, including extraction of 20 difficult-to-treat depression factors using synthetic training data[26], recognition of psychiatric attributes from 150 annotated German mental-state examinations[27], classification of four suicidality-related categories from 500 psychiatric evaluation notes[28], and prediction of acute psychiatric readmission from Danish clinical notes[29]. While these studies establish the feasibility of encoder-based psychiatric NLP, most relied on relatively small annotated corpora or focused on a restricted set of phenotypes, and none addressed span-level extraction of fine-grained psychiatric symptoms at the scale attempted here.

More recently, generative large language models (LLMs) have emerged as powerful alternatives for these tasks. For example, PhenoGPT was developed for identifying Human Phenotype Ontology terms associated with genetic diseases[25]; MentaLLaMA was developed for mental-health analysis of social-media text[30]; and a cross-institutional study evaluated LLMs for extracting 21 social determinants of health from notes at four institutions[31]. Unlike encoder-only models, generative LLMs utilize the Transformer decoder to produce text autoregressively by predicting each token from the preceding text. They are typically larger and more computationally demanding than encoder-only models, often containing billions rather than hundreds of millions of parameters. Nevertheless, they can generalize across applications such as classification, information extraction, summarization, and question answering, often without requiring specialized fine-tuning[32–35]. These capabilities make them promising candidates for flexible symptom detection in clinical text.

Despite these methodological advances, the development of reliable cNLP tools for psychiatric concept extraction remains constrained by several factors. First is the availability of high-quality annotated clinical data, required for model training and evaluation. The limited accessibility of EHR notes and the time-consuming annotation process - requiring manual labeling by expert clinicians - make the assembly of representative datasets difficult[36,37]. This problem is compounded in psychiatry by the fact that many symptoms are infrequent or seldom documented in EHRs (because they may be rare, episodic, inconsistently assessed, or recorded only when clinically salient). This limits the availability of annotated examples and makes their inclusion in model development challenging. Consequently, studies exploring the potential of cNLP models for psychiatric concept extraction typically cover a limited set of concepts and do not target rare symptoms[28,38], rely on non-clinical data[30,39,40], or do not include model generalization evaluations across secondary clinical settings[41,42].

Furthermore, most existing studies leveraging language models for clinical phenotyping focus on English-language settings, with little effort directed toward Spanish-language clinical data, despite Spanish being widely used as a primary language in clinical settings globally[43,44].

Finally, no existing study has evaluated how smaller encoder-based models compare with both manually-developed tNLP and larger generative LLMs for large-scale psychiatric concept extraction under matched development conditions (i.e., developed or fine-tuned using the same data). These comparisons are important because pattern-based methods, encoder-based models, and generative LLMs differ substantially in their development requirements, computational demands, interpretability, and ability to recognize lexically varied expressions. Evaluating these approaches on the same annotations and test data can therefore clarify whether the additional complexity of encoder-based models provides meaningful performance gains over pattern-matching, how relatively small encoder models compare with larger generative LLMs, and which approach may be most appropriate for different psychiatric concepts.

Here, we investigated (i) whether relatively lightweight encoder-only models could accurately detect a broad set of fine-grained psychiatric concepts in Spanish-language EHR notes, (ii) whether weak-label augmentation could improve performance for less frequent concepts, (iii) how well these models generalized to an external clinical setting, and (iv) how their performance compared with tNLP and generative LLMs. To address these aims, we fine-tuned and systematically evaluated three encoder-only Transformer models using clinician-annotated EHR notes from a psychiatric hospital in Colombia affiliated with Misión Origen, a large EHR-linked biobank designed to study psychiatric illnesses in Latin American populations[45].

This study targeted more than 100 clinically meaningful and transdiagnostic psychiatric concepts documented in routine patient care, spanning thought and cognition (e.g., *Delusions*, *Persecutory Ideation*), mood and affect (e.g., *Depressed Mood*, *Hopelessness*), perception (e.g., *Auditory Hallucinations*, *Visual Hallucinations*), and suicidality. These concepts are central to psychiatric diagnosis, risk stratification, and EHR-based research, yet are often absent or under-recorded in coded fields.

To mitigate annotation scarcity for less frequent concepts, we incorporated additional weakly-labeled documents using a tNLP method. Model performance was evaluated primarily on an in-domain test set, and out-of-domain performance was assessed using an independent test set from a second psychiatric facility in Colombia. Finally, the encoder-only models were benchmarked against a tNLP method[46] and a fine-tuned generative LLM[47], each developed using the same original clinician-annotated corpus.

This design distinguishes the present study from prior work by its broad psychiatric symptom-level coverage, Spanish-language setting, evaluation of weighted weak-label augmentation, cross-institutional evaluation, and direct comparison of tNLP, encoder-based, and generative approaches.

## Methods

### Settings and Data

#### Setting

We used EHR documents from Clínica San Juan de Dios Manizales (CSJDM), a psychiatric facility in Colombia serving a population of approximately 1 million individuals, for model fine-tuning and evaluation (Table 1). To examine out-of-domain performance (i.e., performance on data from a clinical setting not used for model fine-tuning), models were also tested on data from the EHR system of Hospital Mental de Antioquia (HOMO), a separate large public psychiatric hospital in Antioquia, Colombia. Both institutions are located in the Paisa region of Colombia, home to the Paisa population, a historically recognized population isolate and the focus of Misión Origen - an EHR-linked biobank in Latin-America developed for the study of psychiatric illnesses[45,48–50].

**Table 1:** Overview of datasets used in this study. This table summarizes the number and composition of annotated documents from each of the two sites. Additionally, we show the composition of the training and test sets, and the sources of strong (clinician-annotated) and weak (tNLP-derived) labels used for model development. The CSJDM dataset provided all training data for fine-tuning, including 1,642 clinician-annotated documents and 12,000 weakly labeled documents used for augmenting low-frequency concepts. The HOMO dataset was used only for external (“out-of-domain”) evaluation.

|  | **Training and Evaluation**<br>**(CSJDM)** | **Out-of-Domain Evaluation**<br>**(HOMO)** |
| --- | --- | --- |
| **Annotated Documents (Total)** | 2,000 | 2,000 |
| **Training Set Documents** | 1,642 | (Not used) |
| **(Strong Labeling)** |  |  |
| **Augmented Training Set Documents (Weak Labeling)** | 12,000 | (Not used) |
| **Test Set Documents** | 358 | 309 |
| **Note Sections Included** | Chief Complaint, Thought Content, Analysis, Current Illness, and Objective | Chief Complaint, Thought Content, Analysis, Affect, and Sensory-Perception |

This study setting is particularly relevant for psychiatric cNLP because these notes contain detailed Spanish-language descriptions of mood, affect, thought content, perception, and behavior that are not adequately represented in structured fields. These symptoms are described through heterogeneous clinical language, including subjective descriptive phrasing, abbreviations, and institution-specific documentation practices across note sections. This makes the study corpus both clinically informative and linguistically challenging, particularly for pattern-based methods that depend on predefined expressions.

#### Annotated Documents (Strong Labeling)

For fine-tuning and testing, we used previously curated datasets of clinical documents, selected and annotated as described in detail in De la Hoz and Frydman-Gani et al.[46]. Briefly, 2,000 EHR documents extracted from the CSJDM EHR note database pertaining to 1,821 patients were annotated by two expert clinicians who identified and labeled all instances of 136 concepts covering a broad spectrum of psychiatric symptoms. These included phenomena related to thought content and cognition, mood and affect, and other behavioral and motor concepts (Figure 1A, Supplementary Table 1). The concept set was adapted from prior psychiatric NLP work in English[16], and augmented with additional site-specific concepts identified through early-stage chart-review, as described in De la Hoz and Frydman-Gani et al.[46]. While 136 concepts were originally annotated, evaluation was restricted to concepts with sufficient representation to enable reliable performance estimates (Supplementary Table 1). This dataset was partitioned into a training set (N=1,642, designated for fine-tuning, Table 1) and a test set (N=358) in a stratified manner, ensuring at least 20% of each concept’s annotated instances appeared in the test set. Inter-annotator agreement on the training set was assessed using Cohen’s Kappa[51], whereas the test set underwent a joint secondary review in which discrepancies were resolved, establishing a gold standard for model evaluation (Figure 1B).

**Figure 1.**
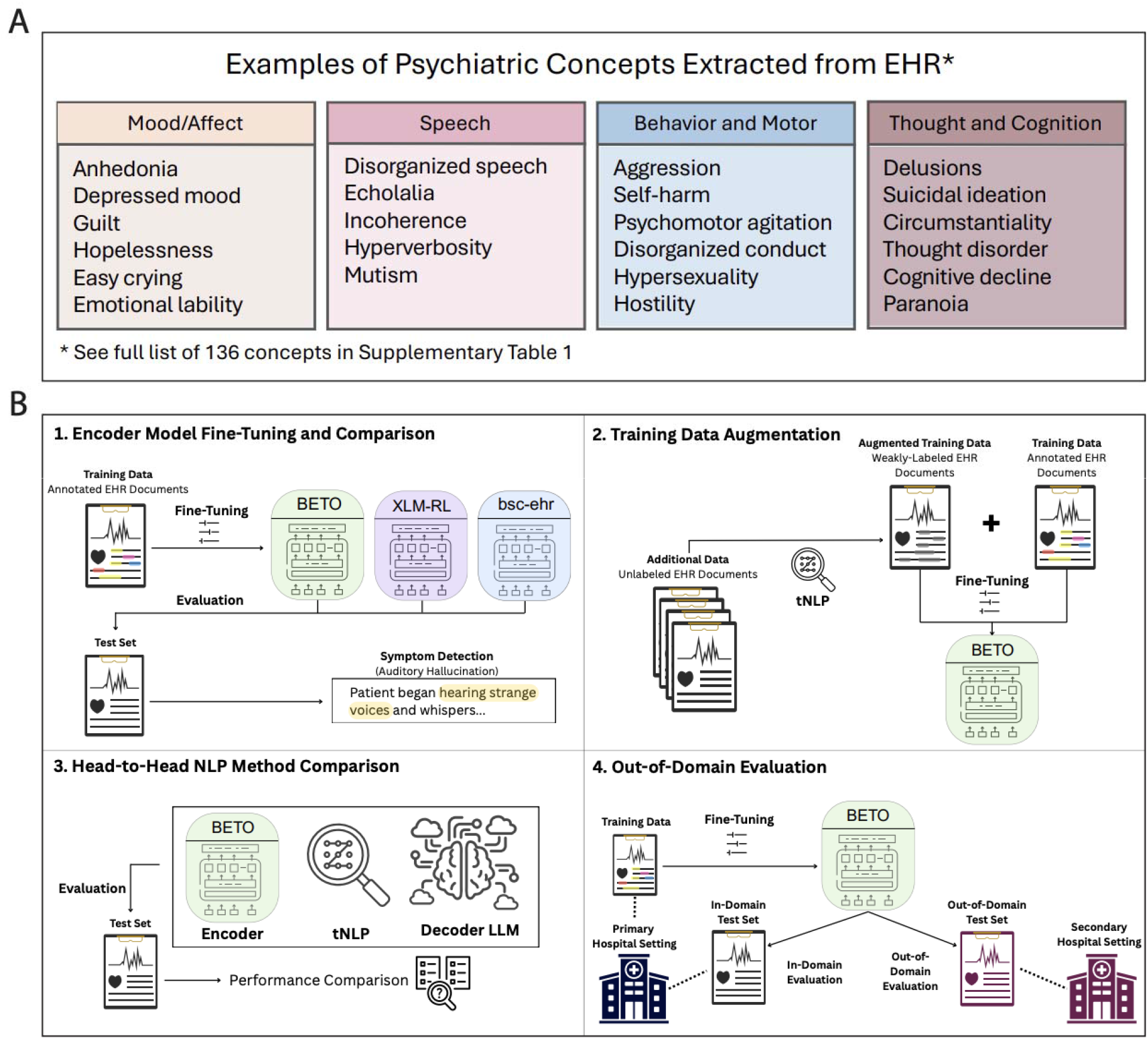
Examples of targeted psychiatric concepts and study workflow. (A) Examples of the psychiatric concepts extracted from Spanish-language EHR notes, spanning domains such as mood and affect, speech, behavior and motor function and thought and cognition. The complete set of 136 concepts annotated is provided in Supplementary Table 1; out of these 136, 110 concepts had sufficient coverage to be included in encoder fine-tuning and evaluation. (B) Overview of the study workflow: (1) Three encoder-based language models - BETO, XLM-RoBERTa-large (“XLM-RL”) and bsc-bio-ehr-es (“bsc-ehr”) - were fine-tuned on 1,642 EHR documents from a psychiatric hospital in Colombia (CSJDM) for the task of psychiatric concept detection, and evaluated on a test set of 358 EHR documents from the same hospital. (2) The effects of augmenting the training data for less frequent concepts (<100 instances in the original training data) were evaluated; the augmented training data consisted of an additional 12,000 documents weakly-labeled by a tNLP system. (3) We compared the performance of the top-performing fine-tuned xbaseline encoder to that of a tNLP method and a top-performing fine-tuned generative LLM (Mistral-small), all developed using the same training data. (4) We performed an out-of-domain evaluation, testing the encoders fine-tuned using data from our primary data source (CSJDM) on data from a second Colombian hospital setting (HOMO), to assess generalizability.

An external test set of 309 documents from HOMO, created using the same workflow[46] and annotated by the same team, was used for out-of-domain-evaluation.

#### Augmented Annotation (Weak Labeling)

For less-frequent concepts (defined here as concepts having fewer than 100 annotated instances in the original training data), we adopted a weak labeling strategy to enrich the CSJDM training set. To assign weak labels, we used a previously developed tNLP method presented in De la Hoz and Frydman-Gani et al.[46], which identifies span-level psychiatric concept mentions in Spanish clinical text using pattern-matching rules. The eligible notes for sampling were recorded between 2016 and 2022 and comprised more than 600,000 notes from nearly 40,000 patients. From these, we randomly sampled without replacement 2,000 unlabeled documents from each of the same six sections of the CSJDM EHR notes used to assemble the original (strong-labeled) fine-tuning data - ‘reason for consultation’, ‘clinical analysis’, ‘current illness’, ‘thought content’, ‘objective’, and ‘analysis’[46] - for a total of 12,000 documents. These sections were originally selected because regex-based prescreening indicated a high concentration of psychiatric symptom mentions. Only documents containing at least 30 words were eligible for sampling, to ensure sufficient textual content for meaningful fine-tuning examples. In addition, we ensured that none of the sampled documents had been included in the original training or test sets.

We then applied the tNLP method to generate weak labels for use in model fine-tuning. To account for differences in reliability between clinician-annotated strong labels and tNLP-derived weak labels, we implemented a weighting scheme in the fine-tuning process. Clinician-annotated documents received a fixed weight of 5, while weakly-labeled documents were weighted proportionally to the precision of the concept-specific tNLP patterns (see Supplementary Note 1 for details). These weights were reflected in the loss function during fine-tuning, ensuring that clinician annotations exerted stronger influence while still including reliable weak labels as augmented examples.

### Model Selection

We selected three open-source encoder-only language models pre-trained on substantial Spanish-language content for fine-tuning and evaluation: BETO, a general-domain Spanish BERT[52]; bsc-bio-ehr-es (hereafter: “bsc-ehr”), a Spanish clinical RoBERTa-based model pre-trained on biomedical and clinical text[53]; and XLM-RoBERTa-large (hereafter: “XLM-RL”), a RoBERTa-based multilingual model[54]. These models represent three complementary strategies: a general-purpose monolingual (BETO), a domain-adapted monolingual encoder (bsc-ehr), and a large-scale multilingual encoder (XLM-RL). This selection allowed us to examine the relative benefits of domain adaptation and cross-lingual transfer for Spanish clinical concept extraction. Additional details about the models and their pre-training corpora are provided in Supplementary Note 2.

### Fine-Tuning

#### Fine-Tuning on Annotated Documents (Strong Labels)

We framed concept extraction as a binary token classification task, predicting whether each token (a basic unit of text, typically consisting of a word, sub-word, or punctuation) was labeled as part of a concept span (I-concept, “inside”) or not (O-concept, “outside”), as suggested by Yang et al.[25]. This choice was motivated by our use of a separate model fine-tuned for each concept, using the annotated CSJDM training set. Because each model only needed to distinguish tokens belonging to a single target concept from all other tokens, overlapping labels (requiring a BIO-style tagging scheme) were not relevant, and the additional boundary label used in BIO-style schemes was less critical than in standard multi-entity NER settings. Given the very limited annotated data available for many concepts, the simpler I/O formulation was selected to reduce sparsity and simplify the learning process.

To construct examples for fine-tuning, the corpus was segmented into 4,050 sentences, with each sentence treated as an independent input sequence paired with token-level annotations. For words split into multiple sub-word tokens, the original word’s label was propagated to all sub-word tokens (see Supplementary Note 3 for details). All three pretrained models - BETO, bsc-ehr, and XLM-RL - were fine-tuned for 10 epochs using the Trainer class from the Hugging Face Transformers library[55] (Supplementary Note 3). To minimize stochastic variations and promote reproducibility, we fixed random seeds across relevant libraries (Python, NumPy, and PyTorch on CPU and GPUs), standardizing randomized operations like training example shuffling, initializations, and drop-out. We also enforced deterministic operations in the NVIDIA CUDA Deep Neural Network library (cuDNN), preventing cuDNN from selecting non-deterministic GPU implementations of operations such as convolutions and matrix multiplications across fine-tuning runs and evaluations.

#### Fine-Tuning on Augmented Annotations (Weak Labels)

To assess whether weak-label augmentation could improve the detection of less-frequent concepts, we created additional weakly-labeled examples for concepts with fewer than 100 annotated instances in the original training data.

The top-performing encoder model was then fine-tuned on this augmented training set (>37,000 sentences total), using the weighting scheme described in (2.1) to distinguish weak- and strong-labeling annotations. Fine-tuning used a weighted cross-entropy loss in which each example’s corresponding loss was scaled by its assigned weight; for additional details and examples, see Supplementary Note 4).

Models fine-tuned exclusively on clinician-annotated data are referred to throughout the manuscript as *baseline* fine-tuned models (e.g., “BETO-Baseline”), whereas models fine-tuned using augmented weak-label annotations are denoted with the suffix *“+WL”* (e.g., “BETO +WL”).

### Evaluation

#### Model Performance Metrics

We evaluated the three encoder-based models on the CSJDM test set, using the clinicians’ annotations as the gold standard. Performance was assessed using precision, recall, and F1. Macro-averaged metrics were calculated by assigning equal weight to each eligible concept. To complement macro-F1 and reduce its sensitivity to variability among rare concepts, we also report micro-F1. To reduce the instability of performance estimates due to low frequency, performance calculations were restricted to concepts with at least three instances in each the original clinician-annotated CSJDM training and test sets, yielding a total of 110 concepts out of the 136 originally annotated.

To quantify uncertainty in the reported encoders’ performance, we performed nonparametric bootstrapping, resampling the CSJDM test set with replacement for 2,000 iterations. For each sample, concept-level precision, recall, and F1 were recomputed for each model. We then calculated bootstrap means and percentile-based 95% confidence intervals.

#### Concept Frequency Sensitivity Analysis

To evaluate how aggregate performance metrics were influenced by rare concepts, we repeated the analyses using progressively stricter minimum-support thresholds. For test-set sensitivity analyses, we included concepts with at least 3, 5, 10, or 20 positive instances in the original CSJDM test set. Similarly, we conducted a sensitivity analysis to assess how aggregate model performance changed with minimal support thresholds in the training data.

#### Comparison to Pattern-Matching tNLP Method and LLM

To contextualize the encoders’ performance, we benchmarked them against two approaches: a tNLP method[46] and a fine-tuned generative LLM[47], Mistral-small. The tNLP patterns had been manually curated using the same CSJDM training set annotations used to fine-tune the encoders in this study, and Mistral-small had also been fine-tuned using this same CSJDM training set. We considered two comparisons: first, matched-training-data comparisons were performed using the baseline encoder (BETO-Baseline), the tNLP method, and Mistral-small, all developed using the same data. Second, weak-label augmentation comparisons were performed by comparing the model trained on the augmented data (BETO +WL, the same encoder architecture fine-tuned on an augmented dataset that additionally included weakly labeled examples), against the tNLP and Mistral-small. These comparisons were designated to assess the effect of weak-label augmentation rather than provide a direct training-matched comparison. Accordingly, comparisons involving BETO +WL and Mistral-small should be interpreted as assessing gap reduction after augmentation rather than parity under equivalent fine-tuning conditions.

For comparison with the tNLP method, model performance was compared on the 88 concepts for which the tNLP method was evaluated in De la Hoz and Frydman-Gani et al.[46] (Supplementary Table 1). In that study, pattern development was restricted to concepts meeting prespecified minimum training-frequency and inter-annotator-agreement criteria (Cohen’s Kappa >=0.6 and training set frequency >=10). For comparison with the generative LLM, performance was evaluated on the 109 concepts for which the LLM was assessed. We selected the fine-tuned Mistral-small generative LLM (24B parameters) for comparison, as it was the smallest among the top-performing LLMs fine-tuned on the CSJDM dataset, as reported in Frydman-Gani et al.[47]. Although 110 phenotypes were evaluated in the present study, the concept *Flight of Ideas* was excluded from the prior LLM evaluation because it did not meet the prespecified minimum of three positive mentions in both the training and test sets after creation of the LLM hyperparameter-tuning validation set. For this reason, the paired encoder-LLM comparison was restricted to the 109 phenotypes shared by both studies.

#### Out-of-Domain Generalizability

The best-performing encoder-based model was evaluated on the HOMO test set to assess out-of-domain performance.

#### Statistical Analysis

To formalize performance comparisons, we conducted paired Wilcoxon signed-rank tests on F1, precision, and recall scores, with false discovery rate (FDR) correction applied within each set of comparisons. This nonparametric test was selected because the same concepts were evaluated across models, yielding paired observations, and because the distributions of concept-level differences were not normally distributed. Because failure to detect a significant difference does not establish equivalence, we also assessed model equivalence using a two one-sided tests (TOST) procedure on paired concept-level differences, with a pre-specified equivalence margin of 0.05.

### Attention Rollout

We performed an illustrative examination of the most attended unigrams (single-word sequences) by looking at attention rollout[56], a post-hoc method measuring global attention used to approximate the relevance of input tokens. We selected three concepts (*Adverse Effects*, *Hallucinations*, and *Suicide Attempt*) to present tokens receiving high model attention. These concepts were selected qualitatively because an encoder substantially outperformed tNLP, particularly in recall, and because they had sufficient frequency and high lexical diversity.

Attention rollout was used as an exploratory interpretability tool rather than for formal global interpretability analysis across all concepts. Rollout was calculated on the test set, and token-level results were averaged to word-level scores as needed, for interpretability.

### Ethical Approval

This study was conducted in compliance with the applicable United States and Colombian regulations and institutional guidelines. The study procedures were reviewed and approved by the University of California, Los Angeles Medical Institutional Review Board 3 (IRB#20-000149, IRB#20-001537, and IRB#16-002084), the Comité de Ética del Instituto de Investigaciones Médicas at Universidad de Antioquia, the Comité de Ética en Investigación de la E.S.E. Hospital Mental de Antioquia, and the Comité de Bioética de Clínica San Juan de Dios Manizales.

Participants provided signed informed consent for participation in the studies approved under IRB#20-000149, IRB#20-001537, and IRB#16-002084. Access to electronic health record data from Clínica San Juan de Dios Manizales was approved by the Comité de Bioética de Clínica San Juan de Dios Manizales with a waiver of informed consent for secondary use of clinical data. For data obtained from Hospital Mental de Antioquia, all participants provided informed consent authorizing access to and secondary analysis of their electronic health record data. All secondary analyses reported in this study were conducted under the ethical approvals and consent provisions described above.

To protect participant privacy and confidentiality, all analyses were conducted using de-identified data stored and processed in secure, access-controlled computing environments. Model development and evaluation were performed on clinical text within these protected environments. Publicly released materials do not include identifiable patient information. The synthetic dataset was generated from previously published, manually reviewed concept annotation spans that did not contain protected health information, and the released synthetic text contains no patient identifiers. No images or supplementary materials contain identifiable individual participants. No identification of individual participants is possible from the manuscript or supplementary material. Participants were not compensated for the analyses described in this manuscript.

## Results

### Model Performance

#### Fine-Tuning on Annotated Documents (Strong Labels)

All fine-tuned encoder-based models were evaluated for detection of 110 concepts (Supplementary Tables 1-4) in the test set. BETO (hereafter: “BETO-Baseline”) and XLM-RL achieved similar performance (macro-F1=0.81, 95% CI= [0.79-0.84]; vs. macro-F1=0.80, 95% CI= [0.79-0.83], respectively; Table 2, Figure 2, Supplementary Tables 2-3). In contrast, bsc-ehr showed significantly poorer performance, with a macro-F1 score of 0.64 (95% CI= [0.63-0.68], *P_FDR_*<.001; Supplementary Table 4).

**Figure 2.**
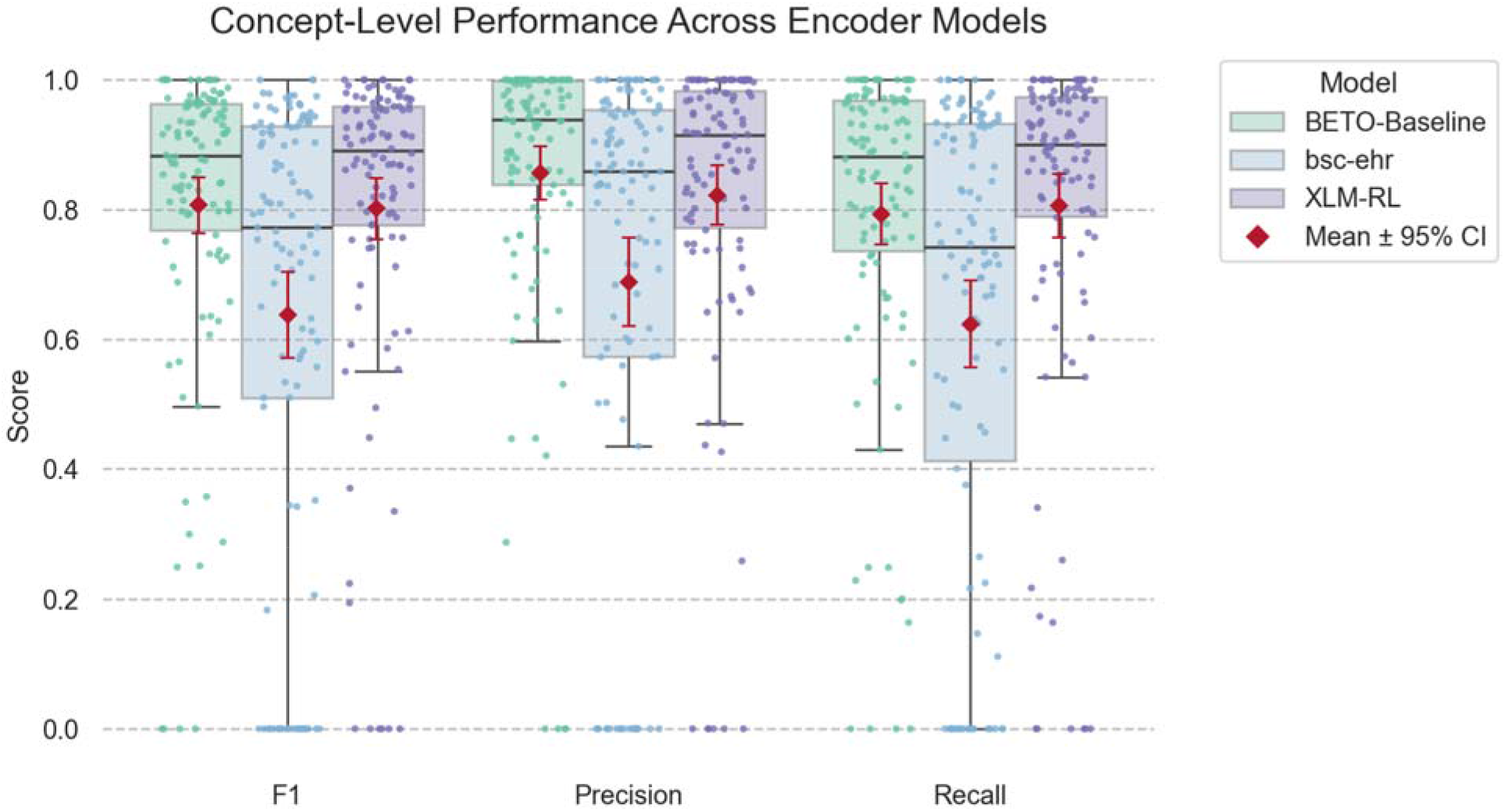
Distribution of concept-level F1, precision, and recall scores for the three fine-tuned encoder-based models: BETO-Baseline (green), bsc-ehr (blue), and XLM-RL (purple). Each point represents performance for a single concept (total concepts=110). Boxplots show the median and interquartile range (IQR) across concepts, with whiskers extending to 1.5×IQR. Red diamonds and error bars indicate the mean and 95% confidence interval.

**Table 2.**
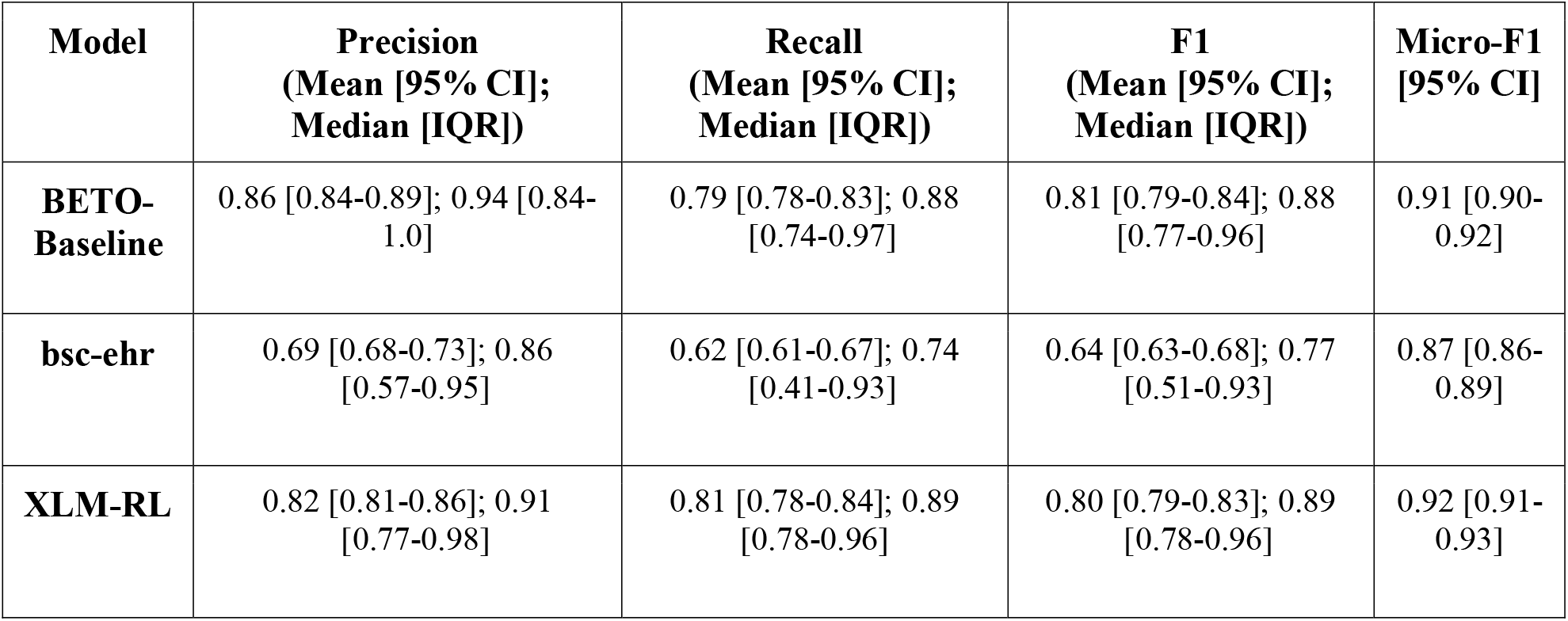
Mean F1, precision, and recall scores [95% CI obtained through bootstrapping] and Median [IQR], across 110 evaluated concepts for the fine-tuned encoder-based models: BETO-Baseline, bsc-ehr, and XLM-RL.

| **Model** | **Precision**<br>**(Mean [95% CI];**<br>**Median [IQR])** | **Recall**<br>**(Mean [95% CI];**<br>**Median [IQR])** | **F1**<br>**(Mean [95% CI];**<br>**Median [IQR])** | **Micro-F1**<br>**[95% CI]** |
| --- | --- | --- | --- | --- |
| **BETO-Baseline** | 0.86 [0.84-0.89]; 0.94 [0.84-1.0] | 0.79 [0.78-0.83]; 0.88 [0.74-0.97] | 0.81 [0.79-0.84]; 0.88 [0.77-0.96] | 0.91 [0.90-0.92] |
| **bsc-ehr** | 0.69 [0.68-0.73]; 0.86 [0.57-0.95] | 0.62 [0.61-0.67]; 0.74 [0.41-0.93] | 0.64 [0.63-0.68]; 0.77 [0.51-0.93] | 0.87 [0.86-0.89] |
| **XLM-RL** | 0.82 [0.81-0.86]; 0.91 [0.77-0.98] | 0.81 [0.78-0.84]; 0.89 [0.78-0.96] | 0.80 [0.79-0.83]; 0.89 [0.78-0.96] | 0.92 [0.91-0.93] |

Model performance was associated with frequency in the training data (Spearman ρ=0.36, *P*<.001 for BETO-Baseline), with diminishing returns at higher frequencies. All models achieved higher macro-F1 scores (0.85-0.91) on concepts with at least 100 instances in the training data, compared to less-frequent concepts (0.54-0.76) and frequent concepts such as *Thoughts of Death*, *Suicidal Ideation*, and *Insomnia* were reliably extracted by all three models.

While BETO-Baseline and XLM-RL maintained relatively stable performance across less-frequent concepts (Supplementary Figures 1 and 2), bsc-ehr showed greater variability (Supplementary Figure 3). For example, *Change in Weight* and *Weight Loss* (with 12 and 10 training instances, respectively) were poorly captured by bsc-ehr but reliably detected by BETO-Baseline and XLM-RL.

The models also exhibited different precision-recall profiles. BETO-Baseline showed higher precision than recall, whereas XLM-RL produced more balanced performance. Bsc-ehr performed substantially worse on both metrics, indicating increases in both missed and false detections rather than a precision/recall trade-off.

#### Fine-Tuning on Augmented Annotations (Weak Labels)

Since BETO-Baseline had the highest macro-F1 when fine-tuned on clinician-annotated data, while smaller than XLM-RL, it was selected for re-fine-tuning with the augmented dataset.

Weak labels were generated for 55 less-frequent concepts (<100 annotated instances) that had available patterns, using the tNLP method developed in De la Hoz & Frydman-Gani et al.[46] (Supplementary Tables 1, 5). Weak labeling substantially increased training support, with 49 of the 55 augmented concepts surpassing 100 training instances in the mixed-label training set.

When evaluated on the CSJDM test set, fine-tuning BETO on the augmented mixed-label dataset (hereafter: “BETO +WL”) resulted in modest improvements compared with BETO-Baseline (delta macro-F1=+0.02, *P_FDR_*=0.02 overall; delta macro-F1=+0.04 on the 55 augmented concepts; Supplementary Table 6).

Particularly significant gains were observed for concepts with limited annotations in the original training set, such as, *Cigarette/Tobacco*, *Perseveration* and *Obsessions* (Figure 3, Supplementary Figures 4-5). These improvements were driven primarily by recall, suggesting that additional training examples improved the model’s concept recognition in unseen text.

**Figure 3.**
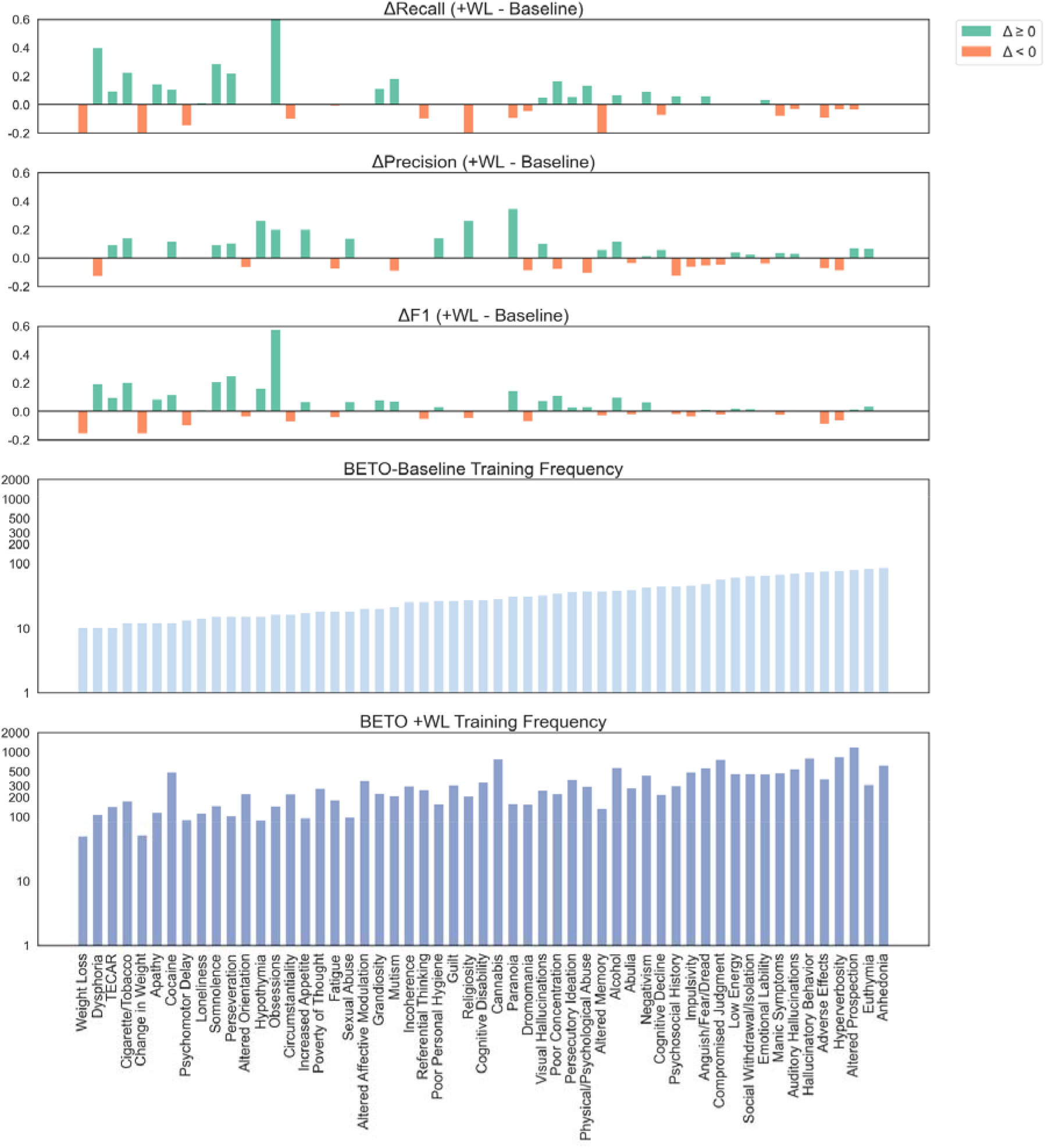
Difference in performance between BETO-Baseline (BETO fine-tuned on the original, strong-labeled training set only) and BETO +WL (BETO fine-tuned on a mixed-labeled training set augmented with weak labels added by the tNLP method) for the 55 less-frequent concepts (<100 instances in the original training set). Performance panels show concept-level changes from BETO-Baseline’s performance in recall (top), precision (center) and F1 (bottom) after incorporating the mixed-label approach. Red bars indicate performance loss (negative change), while green bars indicate performance gains (positive change). Bottom two panels show concept frequencies in the original (light blue) and augmented (dark blue) training sets. Concepts are sorted according to their frequency in the original training set.

Not all concepts benefited to the same extent. Some concepts that were originally difficult for BETO-Baseline to detect, such as *Poor Personal Hygiene* (delta F1=+0.04) and *Physical/Psychological Abuse* (delta F1=+0.04), showed only marginal improvement despite increases in training frequency. Conversely, a few concepts that had performed well under strong-label fine-tuning (e.g., *Weight Loss*) showed worse performance after label augmentation (Supplementary Table 6).

Of the 55 augmented concepts, F1 increased for 30, remained unchanged for 8, and decreased for 17 (Supplementary Table 6). Improvement was inversely associated with baseline F1 (Spearman ρ=-0.56, *P*<.001), and 22/31 of concepts with baseline F1<0.90 improved after augmentation A larger weak-to-strong label ratio was associated with greater recall improvements (ρ=0.34, *P*=.012), whereas pattern precision was not associated with ΔF1. Taken together, these findings suggest that the benefit of augmentation depended more on baseline performance than on concept-specific pattern characteristics.

#### Comparison with tNLP Method and LLM

To contextualize encoder performance, we compared the BETO-based models with a tNLP method and the fine-tuned generative LLM Mistral-small (Table 3, Figure 4, Supplementary Figure 6), a top-performing LMM for this task[47]. These comparisons addressed two different questions: first, matched-training-data comparisons between BETO-Baseline, tNLP, and Mistral-small, all developed using the same training set. Second, we compared the tNLP method and LLM to BETO +WL, which was fine-tuned on an augmented dataset; these comparisons therefore assess the effect of weak-label augmentation, rather than providing a training set-matched comparison.

**Figure 4.**
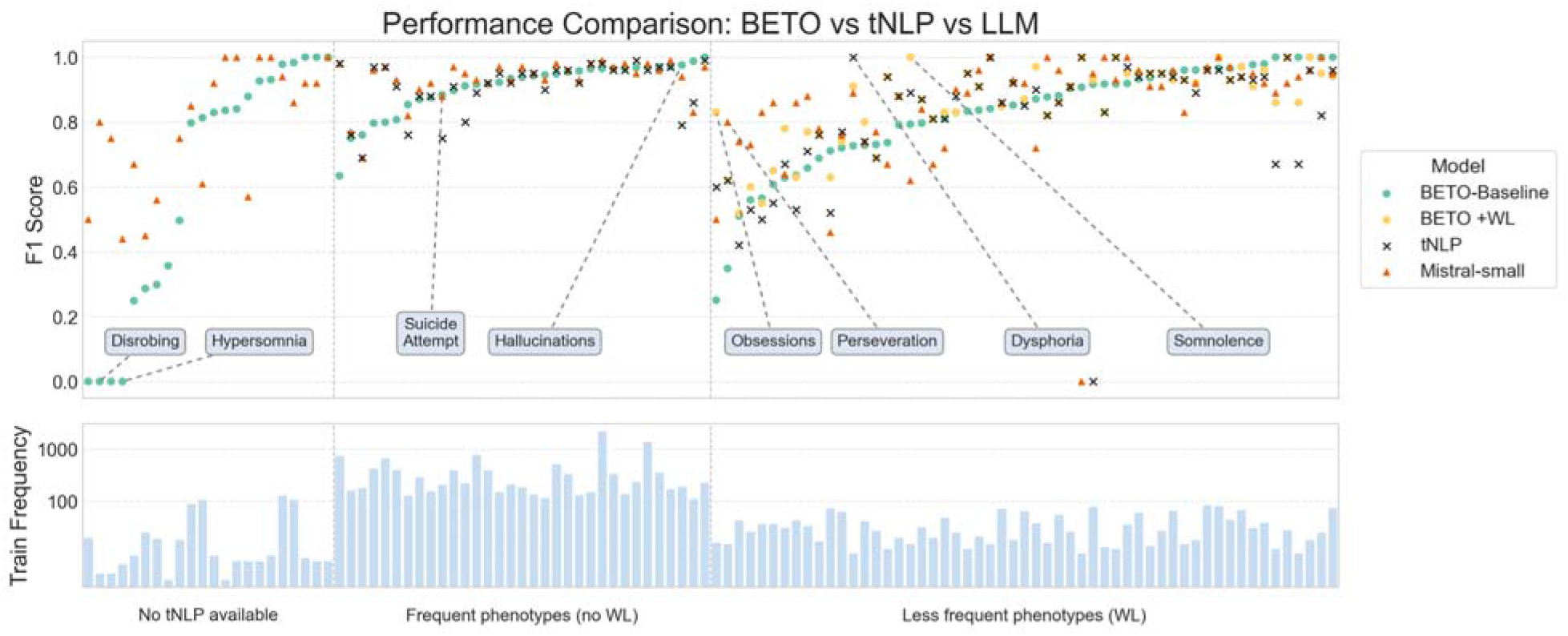
Concept-level performance comparison of BETO-Baseline and BETO +WL against tNLP and a top-performing LLM. The top panel shows concept-level F1 scores for BETO-Baseline (green circles); BETO +WL (yellow circles); tNLP (black crosses); and fine-tuned Mistral-small (orange triangles). Thus, comparisons involving BETO-Baseline, tNLP, and Mistral-small reflect matched-training-data comparisons, whereas comparisons involving BETO +WL reflect the effect of weak-label augmentation rather than a training set-matched benchmark. The bottom panel shows the concepts’ frequency in the fine-tuning data (blue bars, logarithmic scale). The bottom panel shows concept frequency in the original fine-tuning data on a logarithmic scale. Concepts are grouped according to tNLP-pattern availability and weak-label augmentation: no tNLP pattern or augmentation (n=22), tNLP pattern without augmentation because of higher frequency (n=33), and tNLP pattern with weak-label augmentation for less-frequent phenotypes (n=55). Within each group, concepts are ordered by BETO-Baseline F1.

**Table 3.** Performance comparison of fine-tuned encoder-based BETO-Baseline and BETO +WL against a tNLP method and the fine-tuned generative LLM Mistral-small on the held-out CSJDM test set (n=358). The table is divided into two sections: Matched-training-data comparisons show direct comparisons in which BETO-Baseline, tNLP, and Mistral-small were all developed using the original CSJDM training set. Weak-label augmentation comparisons show comparisons involving BETO +WL, the same encoder architecture fine-tuned on an augmented dataset including additional weakly labeled examples; these comparisons assess the effect of weak-label augmentation rather than a training data-matched benchmark. . Because Mistral-small and tNLP were evaluated on 109 and 88 concepts, respectively, comparisons with BETO were restricted to those same shared concept sets. We report mean and median performance across concepts, the mean paired difference (Model 1 - Model 2), the false discovery rate (FDR)-adjusted Wilcoxon signed-rank p-value, and the result of a two one-sided tests (TOST) equivalence analysis for a prespecified margin of ±0.05.

| Performance comparison: paired Wilcoxon signed-rank tests and TOST equivalence analyses for BETO-Baseline and BETO +WL versus tNLP and fine-tuned Mistral-small |  |  |  |  |  |  |  |
| --- | --- | --- | --- | --- | --- | --- | --- |
| Model 1 | Model 2 | Metric | Model 1 Performance<br>(Mean; Median [IQR]) | Model 2 Performance<br>(Mean; Median [IQR]) | Mean paired Δ<br>(Model 1 - Model 2) | Wilcoxon pFDR<br>(*Significance at 0.05) | TOST equivalence within ±0.05 |
| Matched-Training Data Comparisons |  |  |  |  |  |  |  |
| BETO-Baseline | Mistral-small | F1 | 0.81; 0.88 [0.79-0.96] | 0.86; 0.92 [0.80–0.96] | -0.05 | 9.66×10<sup>-3</sup> (*) | No |
|  |  | Precision | 0.86; 0.94 [0.84-1.0] | 0.90; 0.95 [0.86–1.0] | -0.04 | 0.164 | No |
|  |  | Recall | 0.80; 0.88 [0.75-0.97] | 0.85; 0.93 [0.78–0.99] | -0.05 | 5.81×10<sup>-3</sup> (*) | No |
| BETO-Baseline | tNLP | F1 | 0.86; 0.91 [0.80–0.96] | 0.85; 0.91 [0.80–0.96] | 0.01 | 0.948 | Yes |
|  |  | Precision | 0.90; 0.94 [0.86–1.0] | 0.91; 0.96 [0.88–1.0] | -0.01 | 0.164 | Yes |
|  |  | Recall | 0.85; 0.90 [0.79–0.97] | 0.82; 0.90 [0.75–0.96] | 0.03 | 0.365 | No |
| Weak-Label Augmentation Comparisons |  |  |  |  |  |  |  |
| BETO +WL | Mistral- | F1 | 0.83; 0.91 [0.81–0.96] | 0.86; 0.92 [0.80–0.96] | -0.03 | 7.87×10<sup>-2</sup> | No |
|  | small | Precision | 0.87; 0.95 [0.85–1] | 0.90; 0.95 [0.86–1.0] | -0.03 | 0.766 | No |
| | | Recall | 0.81; 0.89 [0.75–0.97] | 0.85; 0.93 [0.78–0.99] | -0.04 | $1.01 \times 10^{-2}$ (*) | No |
| BETO +WL | tNLP | F1 | 0.88; 0.92 [0.83–0.96] | 0.85; 0.91 [0.80–0.96] | 0.03 | $9.66 \times 10^{-3}$ (*) | No |
|  |  | Precision | 0.91; 0.95 [0.88–1.0] | 0.91; 0.96 [0.88–1.0] | 0.01 | 0.783 | Yes |
| | | Recall | 0.86; 0.91 [0.79–0.97] | 0.82; 0.90 [0.75–0.96] | 0.04 | $4.89 \times 10^{-3}$ (*) | No |

Across the 109 concepts evaluated for fine-tuned Mistral-small, Mistral-small significantly outperformed BETO-Baseline in F1 and recall (macro-F1=0.86 vs. 0.81, *P_FDR_*=.0096; Table 3, Supplementary Tables 7-8), whereas precision did not significantly differ; formal equivalence was not established for any metric. In contrast, BETO-Baseline and tNLP did not significantly differ in F1, precision, or recall by Wilcoxon testing (macro-F1=0.86 vs. 0.85; *P_FDR_*=0.948); TOST supported equivalence for F1 and precision within the prespecified ±0.05 margin, but not for recall.

We next evaluated the effect of weak-label augmentation by comparing BETO +WL with tNLP and Mistral-small. Relative to Mistral-small, BETO +WL reduced the performance gap across all metrics, and Wilcoxon tests detected no significant differences in F1 or precision (macro-F1=0.83 vs. 0.86; *P_FDR_*=0.13); However, recall remained significantly lower (*P_FDR_*=0.01), and equivalence was not established for any metric.

In contrast, BETO +WL significantly outperformed the tNLP method in F1 (macro-F1=0.88 vs. macro-F1=0.85; *P_FDR_*<.001) and recall (mean recall=0.86 vs. 0.82; *P_FDR_*<.001), while precision remained equivalent within the prespecified ±0.05 margin (Supplementary Table 8).

Concept-level analyses further revealed predictable patterns. Among n concepts not included in the tNLP evaluation (including many low-frequency concepts), Mistral-small generally outperformed BETO-Baseline, with the latter failing to detect several low-frequency concepts such as *Disrobing* and *Hypersomnia* (F1=0, Figure 4, Supplementary Table 7). BETO-Baseline exceeded tNLP on several concepts such as *Suicide Attempt* (ΔF1=+0.13) and *Hallucinations* (ΔF1=+0.19). Among the 55 augmented concepts, BETO +WL maintained F1>0.5 and outperformed tNLP and Mistral-small on several symptoms (e.g., *Obsessions*, *Somnolence*), while trailing on others (*Perseveration*, *Dysphoria*).

To qualitatively assess the linguistic variability better captured by encoders over tNLP, we reviewed mentions for *Weight Loss*, *Religiosity*, and *Suicide Attempt*-three concepts for which an encoder achieved significantly higher recall. Representative mentions detected by an encoder but missed by tNLP included colloquial descriptions (“*m estoy adelgazando” [sic]*; “I am getting thinner”); morphological variations (“*místico religiosas”;* “mystical-religious”); and indirect expressions (“*me creo dios”*; “I believe I am God”; Supplementary Table 9). These examples illustrate the encoders’ ability to detect semantically related expressions not represented in predefined patterns.

#### Concept Frequency Sensitivity Analysis

To show how the aggregate results were influenced by concepts near the minimum inclusion threshold, we re-calculated F1 using progressively stricter training- and test-set support thresholds (Supplementary Figure 7A, Supplementary Table 10). Macro-F1 increased as minimum test-set support rose from 3 to 20 instances, with the largest gain for bsc-ehr (0.64 to 0.80) and smaller gains for BETO-Baseline, BETO +WL, XLM-RL, and Mistral-small (Supplementary Figure 7A; Supplementary Table 10). In the 3-9-instance stratum, bsc-ehr performed particularly poorly (mean F1=0.21), whereas the other models ranged from 0.65 to 0.82. Thus, low support increased variability but did not primarily drive the aggregate performance of the BETO-based models. These findings indicate that low test-set support contributed to performance variability, but did not primarily drive the aggregate performance of the BETO-based models.

Increasing minimum training support produced a similar pattern, with the greatest improvement for bsc-ehr and more modest gains for the other models (Supplementary Figure 7B), suggesting that limited training data contributed disproportionately to bsc-ehr’s poorer performance.

### Out-of-Domain Testing

To assess cross-institutional generalization, we evaluated BETO +WL on the HOMO test set. Performance was compared for concepts with at least three instances in the CSJDM and HOMO test sets (N=98). Macro-F1 decreased from 0.84 on the CSJDM test set to 0.78 on the HOMO test set (Supplementary Figure 8, Supplementary Table 11), a statistically significant decrease (*P*=.0073). For comparison, the LLM exhibited a similar decline rate (0.86 to 0.81) under the same out-of-domain evaluation[47]. The decline was larger for recall than for precision (Δrecall=-0.06 versus Δprecision=-0.03), suggesting that the reduction in performance primarily reflected an increase in missed gold-standard mentions rather than a comparable loss of precision.

Notably, many concepts (35) showed improved detection performance in the HOMO test set (Supplementary Figure 9, Supplementary Table 11). For instance, *Other Addictions* (delta F1=+0.43) and *Cognitive Decline* (delta F1=+0.30) showed higher F1 when evaluated on HOMO, due to recurring expression patterns shared between the fine-tuning data and the HOMO test set. However, performance was more variable on HOMO overall (Supplementary Figures 8, 9), and several concepts experienced drops in performance despite strong in-domain results, including *Psychomotor Delay* (delta F1=-0.82) and *Elevated Mood* (delta F1=-0.88).

Differences in support partially explained cross-site variability - while change in F1 across sites was not significantly associated with CSJDM training frequency alone (Spearman ρ=-0.05, *P*=.60) or HOMO test-set support alone (ρ=0.11, *P*=.26), larger proportional shifts in normalized support between the two test sets were modestly associated with greater declines in F1 (ρ=-0.25, *P*=.012). Lower HOMO support was associated with wider bootstrap F1 confidence intervals (ρ=-0.40, *P*<.001), indicating greater uncertainty for concepts with limited out-of-domain representation. Support differences did not fully account for the observed heterogeneity: substantial declines were also observed for *Referential Thinking*, *Negative Symptoms*, and *Psychomotor Alterations* despite comparable or greater support at HOMO. These residual differences may reflect cross-institutional variation in note composition, documentation practices, or linguistic expression.

### Attention Rollout

We examined the attention rollout scores for *Adverse Effects*, *Hallucinations*, and *Suicide Attempt* to identify the most attended unigrams in the test-set for BETO +WL (Figure 5). These concepts were chosen to qualitatively show encoder behavior in concepts with high lexical variability that were more challenging for tNLP, as all showed higher performance with fine-tuned BETO-Baseline and BETO +WL (+0.11, +0.19, +0.13 in F1, respectively) and diverse annotations. While not a direct measure of importance, these results may provide insight into BETO’s stronger performance: the most attended unigrams target a varied range of symptom manifestations (e.g., diverse psychiatric and somatic adverse effects) that would be challenging to capture using pattern matching. Attention rollout is shown here as an illustrative, example-based analysis rather than a formal global interpretability analysis across all concepts.

**Figure 5.**
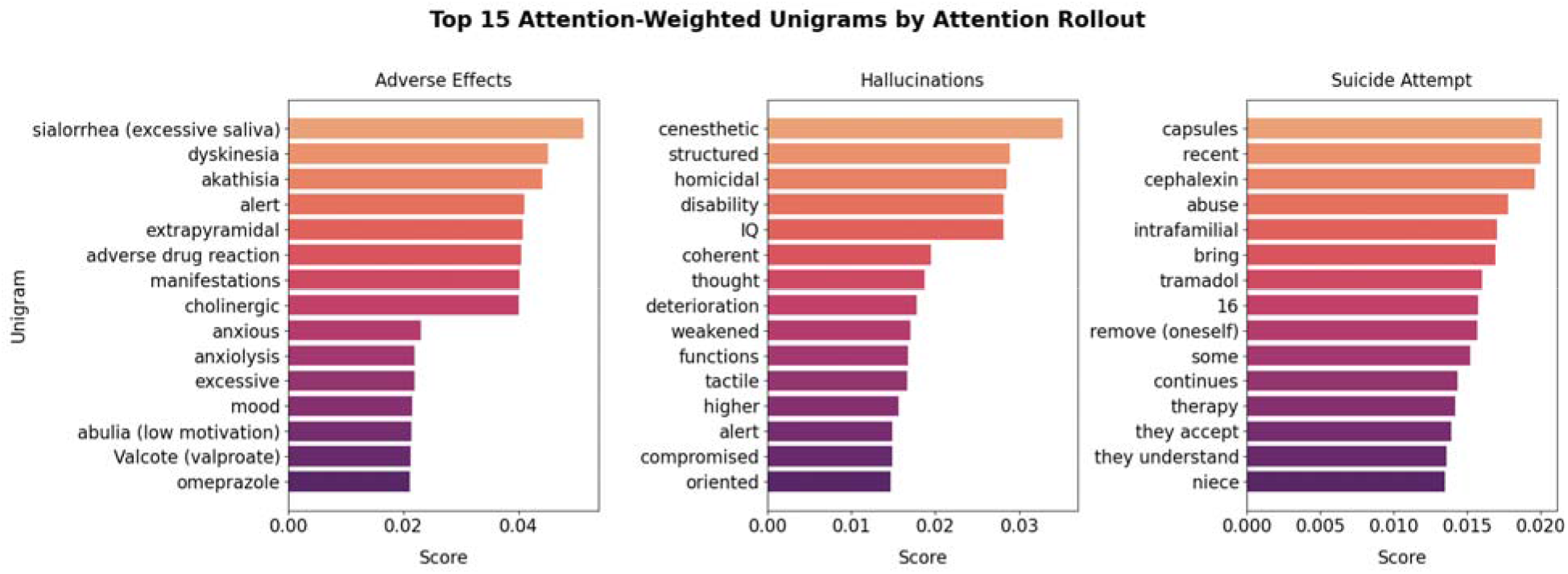
Illustrative attention rollout examples for *Adverse Effects*, *Hallucinations*, and *Suicide Attempt* in fine-tuned BETO +WL. Attention rollout results showing the top 15 most attended unigrams (translated to English from Spanish) for *Adverse Effects* (left), *Hallucinations* (middle) and *Suicide Attempt* (right) for fine-tuned BETO +WL when applied to the test set. These three concepts were selected qualitatively because encoder-based BETO showed substantially higher F1 than the tNLP method on them, with improvements driven largely by higher recall, and because they exhibited diverse expressions in the annotated notes.

## Discussion

### Principal Findings

This study aimed to evaluate encoder-only language models for detecting a broad set of psychiatric concepts in Spanish clinical text. We also assessed whether a weighted weak-label augmentation could improve performance for less-frequent concepts, how well these models generalized to an external clinical setting, and how they compared with an existing tNLP baseline and generative LLMs fine-tuned on the same data.

We found that BETO and XLM-RL achieved the strongest overall and most consistent performance, while the clinical-domain bsc-ehr performed substantially worse. Incorporating weak labels into BETO’s fine-tuning offered modest overall performance improvements, particularly for the rarest concepts, driven primarily by gains in recall without loss of precision.

BETO-Baseline was equivalent to the tNLP system as measured by F1 and precision, whereas Mistral-small significantly outperformed BETO-Baseline in F1 and recall. After weak-label augmentation, BETO +WL significantly outperformed tNLP in F1 and recall while maintaining equivalent precision. Weak labeling also narrowed the performance gap with Mistral-small, although equivalence to Mistral-small was not established. Of note, because BETO +WL was trained on additional weakly-labeled data, this comparison reflects the effect of augmentation rather than a training set-matched comparison. BETO +WL also maintained reasonably strong out-of-domain performance on an external dataset from a second psychiatric facility, although with greater concept-level variability than in the in-domain evaluation.

### Interpretation and Comparison with Prior Work

This study makes several contributions to advancing cNLP capabilities for psychiatry.

First, it provides a systematic evaluation of different encoder-based models for extracting a broad set of psychiatric concepts from Spanish-language clinical notes, while explicitly evaluating performance on a subset with sufficient support for reliable performance estimation. By targeting 110 psychiatric concepts across multiple encoders, this study expands evidence for the feasibility of using encoder-only models for broad psychiatric concept extraction from real-world Spanish EHR text.

In line with prior work[57], our comparisons suggest that domain-specific Spanish encoders do not necessarily outperform general multilingual models on downstream psychiatric concept extraction, a target task that is highly specialized. Despite its biomedical and clinical pre-training, bsc-ehr achieved lower precision, recall, and F1 than BETO-Baseline and XLM-RL, with frequent detection failures for low-support concepts. Of note, most of bsc-ehr’s pre-training corpus consisted of biomedical rather than EHR text, and its clinical component was not specifically derived from psychiatric records[53]. Moreover, its original evaluation tasks primarily involved relatively standardized biomedical entities, whereas psychiatric symptoms in the present data were often expressed through heterogeneous, indirect, or colloquial language.

General-purpose Spanish or multilingual pre-training may therefore have provided broader lexical and contextual coverage for this task. Consistent with this interpretation, the performance gap narrowed when analyses were restricted to concepts with greater training support, suggesting that bsc-ehr may require more task-specific examples to adapt to psychiatric language.

Several concepts proved particularly challenging to extract with tNLP, owing to how they are expressed in this clinical setting. *Thoughts of Death*, *Suicidal Ideation*, *Suicide Attempt*, *Delusions* and *Religiosity*, and *Auditory Hallucinations*, while central to psychiatric diagnosis and severity assessment, are frequently described in free-text using variable, colloquial, or indirect language, and context-specific or culturally-dependent phrasing (e.g., *Suicide Attempt*: “intentó tirarse de un puente” [“tried to throw themselves off a bridge”], *Auditory Hallucinations*: “escucho una voz que dice…” [“I hear a voice that says…”]). Encoder-based models were often able to capture these diverse expressions-they achieved higher recall than tNLP for concepts like *Religiosity* and *Suicide Attempt*, suggesting better capture of non-standard or context-dependent expressions not covered by the predefined patterns.

Our comparisons with tNLP and an LLM, which offer one of the first direct comparisons of these three approaches for psychiatric concept detection, highlight the practical utility of encoder-based models for Spanish psychiatric cNLP and help clarify where encoder-based models sit relative to other options. Pattern-based approaches are attractive because of their speed, lightness and interpretability, but they can be limited by lexical rigidity. In contrast, encoder-based language models can learn more flexible contextual representations - that is, context-sensitive representations not restricted to predefined phrases, allowing the recognition of semantically similar and varied expressions of the same symptoms. In this study, BETO-Baseline was equivalent to tNLP in F1 within the prespecified ±0.05 margin, and weak-label augmentation allowed BETO +WL to surpass tNLP in F1 and recall while maintaining equivalent precision. This pattern shows that encoder-based models can preserve the precision needed for psychiatric concept extraction while improving recall of the varied and indirect language found in psychiatric notes. This interpretation is further supported by the attention rollout illustrations, which highlight a varied set of expressions that would be difficult to capture exhaustively using fixed rules.

The comparison with the generative LLM is also notable: Despite being orders of magnitude smaller than Mistral-small (110M vs 24B parameters) and more efficient to fine-tune and apply, the performance of BETO-Baseline remained reasonably close to the larger model under matched fine-tuning conditions, and weak-label augmentation reduced this gap further. However, performance equivalence to Mistral-small could not be established within the prespecified ±0.05 margin. Of note, because BETO +WL was trained on additional weakly labeled data, this reduction should be interpreted as gap reduction after augmentation rather than parity under equivalent training sets. At the same time, the reduction in this gap suggests that smaller encoder-based models can still achieve strong performance on this supervised psychiatric concept extraction task while requiring substantially fewer computational resources. This is particularly important for real-world EHR repositories, where scalability, cost, and deployment feasibility are central considerations. Rather than replacing encoder models, the emergence of generative LLMs reinforces the need to identify which model class provides the best performance-cost tradeoff for a given clinical cNLP task. Additionally, these findings suggest that weakly supervised augmentation may be especially valuable in low-resource clinical NLP settings, where large manually annotated corpora are difficult to obtain and training or applying very large generative models on vast volumes of notes may be financially and computationally unfeasible. In such settings, augmenting smaller encoder-based models with carefully weighted weak labels may provide a practical way to improve performance while lowering barriers to developing useful psychiatric phenotyping tools. This could be particularly important for reducing global disparities in clinical NLP research, which has historically been concentrated in English-language and higher-resource environments.

### Cross-Site Generalizability

External evaluation showed that BETO +WL achieved reasonably strong performance on an independent dataset from a second psychiatric hospital, supporting partial transportability across institutions. However, performance was more variable compared to the in-domain evaluation. These differences are likely due to differences in concept frequency, patient populations, expressions and documentation practices across institutions, even within the same country and medical specialty.

### Implications for Spanish-Language Psychiatric cNLP

By focusing on Spanish EHR notes, this study addresses a pressing gap in cNLP research. Spanish is the native language of over 450 million people worldwide, and the primary language used in clinical settings across Latin America and Spain, yet is under-represented in cNLP resources. To our knowledge, no directly comparable Spanish-language datasets combine expert clinician annotations for a large number of psychiatric concepts, nor do they combine data from multiple hospital systems. This breadth permits evaluation across diverse and rare symptoms, many of which appear primarily in free-text notes. The inclusion of data from HOMO also permits assessment of generalization within the Paisa region.

Our findings demonstrate the applicability of encoder-based models for concept extraction from real-world Spanish clinical text, even for concepts characterized by variable wording, having practical implications for psychiatric research in Spanish-speaking populations. With appropriate post-processing, validation, and aggregation across documents, encoder-based models such as those evaluated here may enable large-scale extraction of symptom-level information from Spanish EHR repositories containing hundreds of thousands of documents, supporting cohort construction, treatment-response studies, and discovery of clinically meaningful subgroups. The extracted mentions provide a foundation for reducing manual chart annotation efforts and developing phenotypic infrastructure for large-scale EHR-based initiatives such as Misión Origen. Moreover, because these models are smaller and less computationally demanding than many generative LLMs, they are more feasible in settings with limited computational resources or stricter infrastructure constraints.

### Limitations

This study has several limitations that should be considered when interpreting the findings. First, model fine-tuning was conducted without a dedicated validation set for hyperparameter fine-tuning, as partitioning the already limited annotated corpus would have further reduced the number of rare concepts available for fine-tuning. This choice may have limited peak performance: optimization of parameters such as the learning rate, batch size, and number of epochs could potentially have improved performance. Moreover, because the same fine-tuning configuration was applied across encoders, it may not have been equally optimal for all models; Nevertheless, the BETO-based models achieved high performances under these conditions, suggesting the robustness of encoder models for this task. Future work should explore hyperparameter optimization to maximize the potential of encoder-based models, and clarify whether part of bsc-ehr’s underperformance reflected greater sensitivity to fine-tuning settings.

Second, while the annotated dataset was carefully curated and extensive, many concepts were infrequently represented, which contributed to variability in model performance. Augmenting the fine-tuning data with weak labels partially mitigated this limitation; However, because the weak labels rely on predefined patterns, they may not capture synonyms, spelling variants, and other variability not represented in the annotations, limiting the performance on lexically variable concepts. Future work could examine more advanced weak supervision strategies to better harness weak labels while minimizing error propagation. Importantly, the breadth of the annotation set should be distinguished from reliable model performance across concepts: while 136 concepts were originally annotated, only 110 met minimum support criteria for evaluation, and among those, performance on the less-frequent concepts remained most variable. We nevertheless retained rare concepts because they include many clinically valuable psychiatric phenomena, and excluding them would have narrowed the clinical scope of the evaluation. Accordingly, aggregate results should be interpreted alongside concept-level performance and support, and model selection should consider the specific concepts of interest.

Third, both test sets used in this study originated from hospitals within the Paisa region of Colombia, which may constrain the generalizability of the findings to other Spanish-speaking settings. Clinical Spanish may vary across regions, and documentation practices may differ across institutions and note types. Future work should evaluate these models across additional Spanish-speaking settings and examine whether multi-institutional fine-tuning improves generalizability.

Fourth, we did not evaluate whether weak-label augmentation would provide similar benefits for the generative LLM. Although BETO +WL reduced the performance gap with Mistral-small, this comparison reflects the effect of augmenting the training data rather than a strictly matched comparison between modalities for this task, because Mistral-small could not be re-fine-tuned on the augmented dataset. Therefore, while our findings support weak-label augmentation as a practical strategy for improving encoder-based psychiatric concept extraction, they do not establish whether similar gains would be observed for generative LLMs. Future work should directly test weak-label augmentation in both model classes to determine whether its benefits are more pronounced for smaller encoders in low-resource annotation settings.

## Conclusions

This study directly compared encoder-based transformers with both traditional rule-based NLP and generative LLMs for extensive psychiatric concept extraction across a large set of clinically relevant symptoms. A lightweight, general-domain encoder demonstrated strong suitability for cNLP in psychiatry. More broadly, these findings support the development of scalable, efficient, and reliable cNLP tools to advance psychiatric research and patient care in Spanish-speaking populations. As cNLP increasingly incorporates large generative models, this study highlights the continued value of smaller encoder-based models as practical, resource-efficient tools for real-world psychiatric research. Developing and validating such tools across diverse Spanish-speaking clinical settings will be essential for setting up more globally inclusive psychiatric studies, and advancing precision psychiatry in underrepresented populations.

## Acknowledgments

During manuscript revision, ChatGPT 5 (OpenAI) was used to assist with language editing and sentence flow of author-written text (proofreading and language editing). The tool was not used to generate scientific analyses, hypotheses, results, references, figures, or conclusions. All text was reviewed, edited, verified, and approved by the authors, who take full responsibility for the final content of the manuscript.

## Compliance Statement

All procedures were performed in compliance with Colombian and United States laws and institutional guidelines and have been approved by the Institutional Review Board (IRB), Medical Institutional Review Board 3, at UCLA (IRB#20-000149, IRB#20-001537 and IRB#16-002084), the Comité de Ética del Instituto de Investigaciones Médicas at Universidad de Antioquia (UdeA), the Comité de Ética en Investigación de la E.S.E at HOMO, and the Comité de Bioética de Clínica San Juan de Dios at CSJDM. Participants provided signed informed consent to be part of studies IRB#20-000149, IRB#20-001537 and IRB#16-002084; in addition, a waiver of consent and IRB approval by Comité de Bioética de Clínica San Juan de Dios at CSJDM was granted to access all EHR data. All privacy rights of human subjects have been observed.

## Data Availability Statement

The full annotated corpus used to evaluate and fine-tune all models consists of clinical notes containing patient data, and cannot be shared publicly; however, the specific spans annotated for all concepts do not contain private health information, and were previously published in De la Hoz & Frydman-Gani et al.[46]. The tNLP method’s patterns have also been made public under the same study[46], available under: https://github.com/clarafrydman/Spanish_Psych_Phenotyping.

## Author Contributions Statement

XX and CFG: conceptualization, methodology, analysis, writing of original draft; AA: data curation, methodology, analysis; MPV and JDLM: methodology, annotation of clinical notes; JVE: project administration; MC: methodology; NBF: conceptualization, methodology, supervision, writing of original draft; CLJ: methodology, data curation, supervision; LMOL: conceptualization, methodology, supervision, writing of original draft. All authors read, edited and approved the final manuscript.

