## Supplementary Figures and Notes for "Large-Scale Psychiatric Concept Extraction from Electronic Health Records: A Comparative Study of Encoder-Based Language Models"

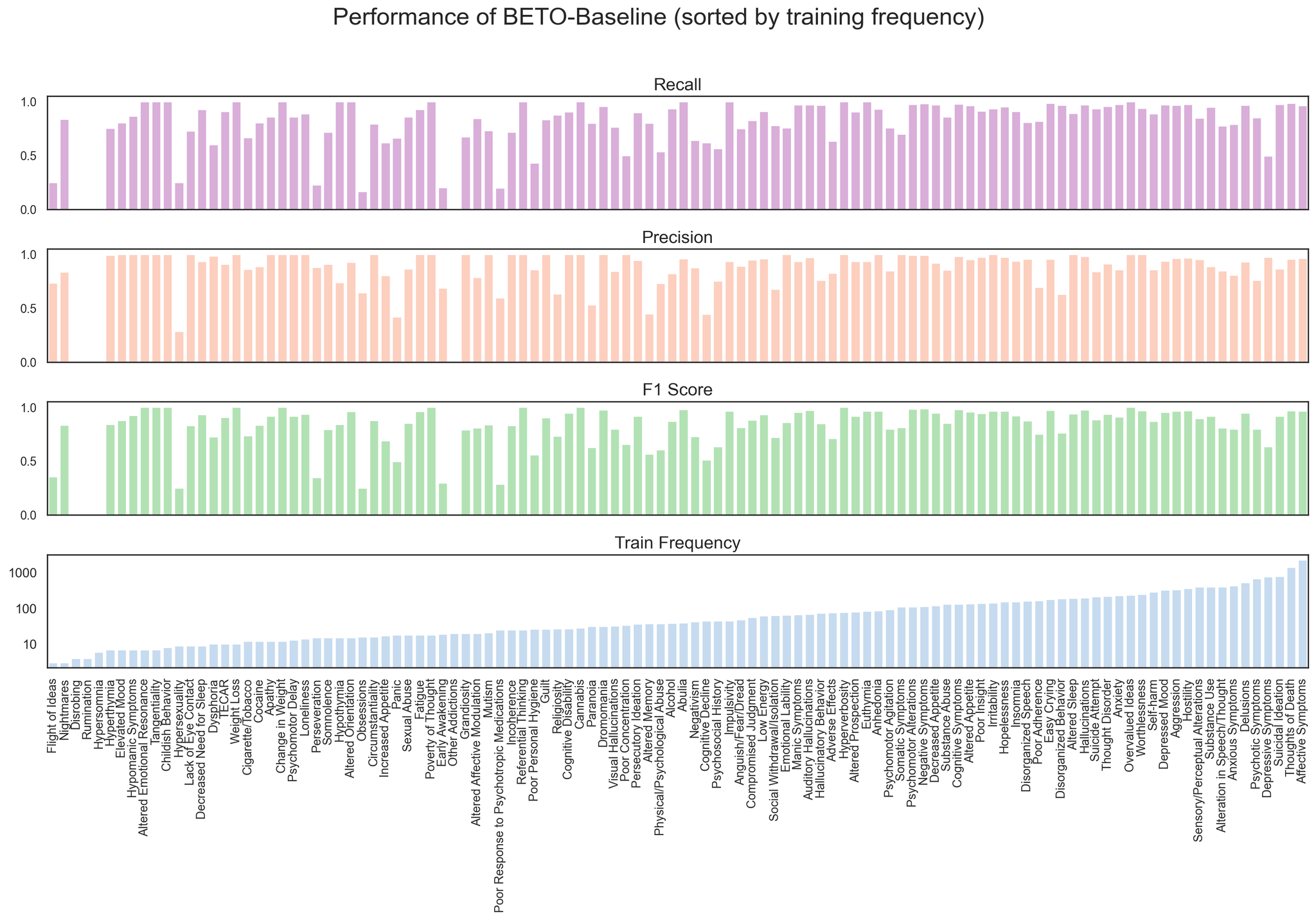

**Supplementary Figure 1**. Per-concept performance of BETO-Baseline model fine-tuned on the CSJDM training set, evaluated on the CSJDM test set (n=358 documents, n=110 concepts). The three performance panels show per-concept recall (top, pink), precision (middle, orange), and F1 score (bottom, green). The bottom panel shows the training set frequency for each concept on a log_10_ scale (blue). Concepts are ordered by their frequency in the training set (increasing left to right).

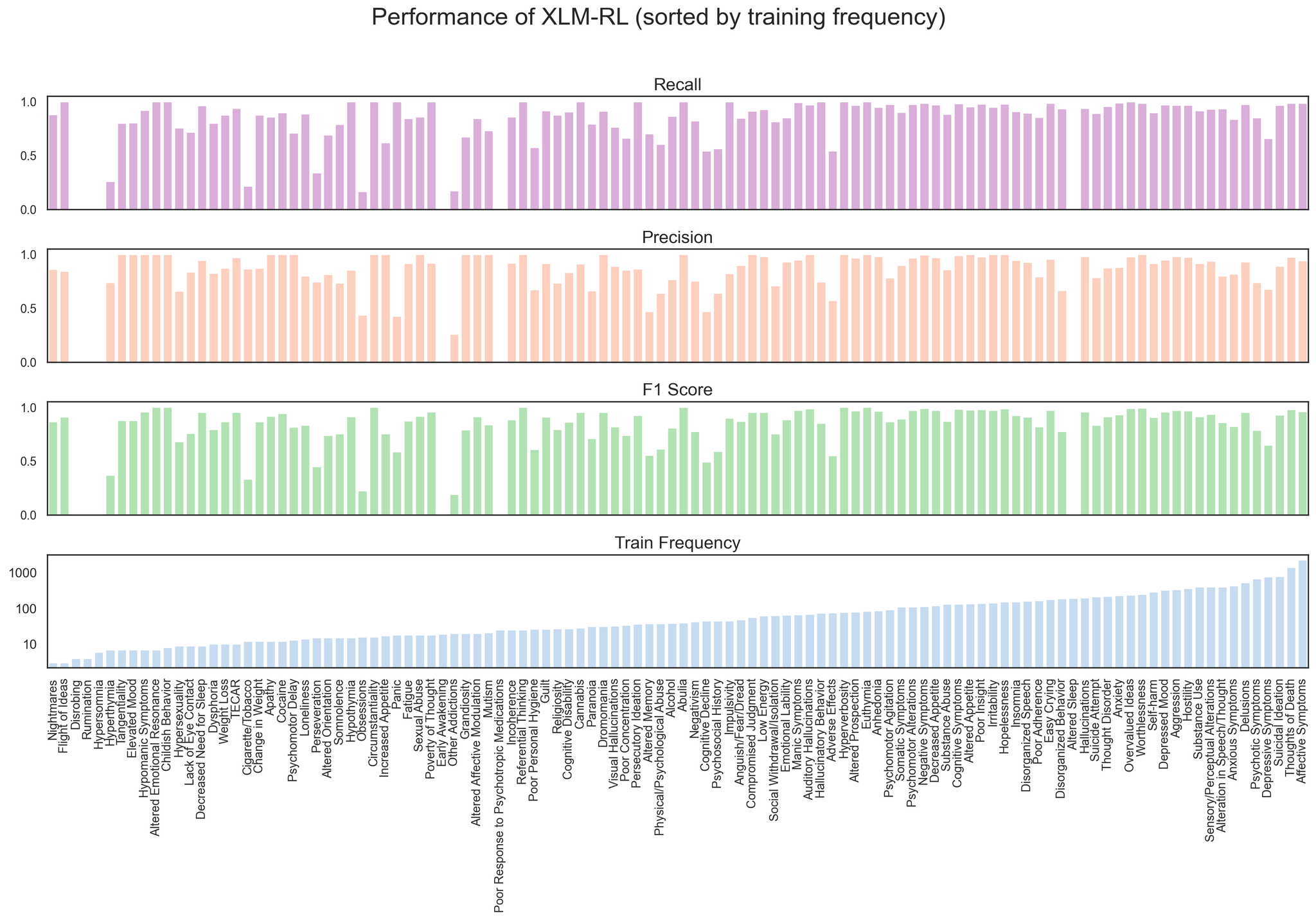

**Supplementary Figure 2**. Per-concept performance of XLM-RL fine-tuned on the CSJDM training set, evaluated on the CSJDM test set (n=358 documents, n=110 concepts). The three performance panels show per-concept recall (top, pink), precision (middle, orange), and F1 score (bottom, green). The bottom panel shows the training set frequency for each concept on a log_10_ scale (blue). Concepts are ordered by their training frequency (increasing left to right).

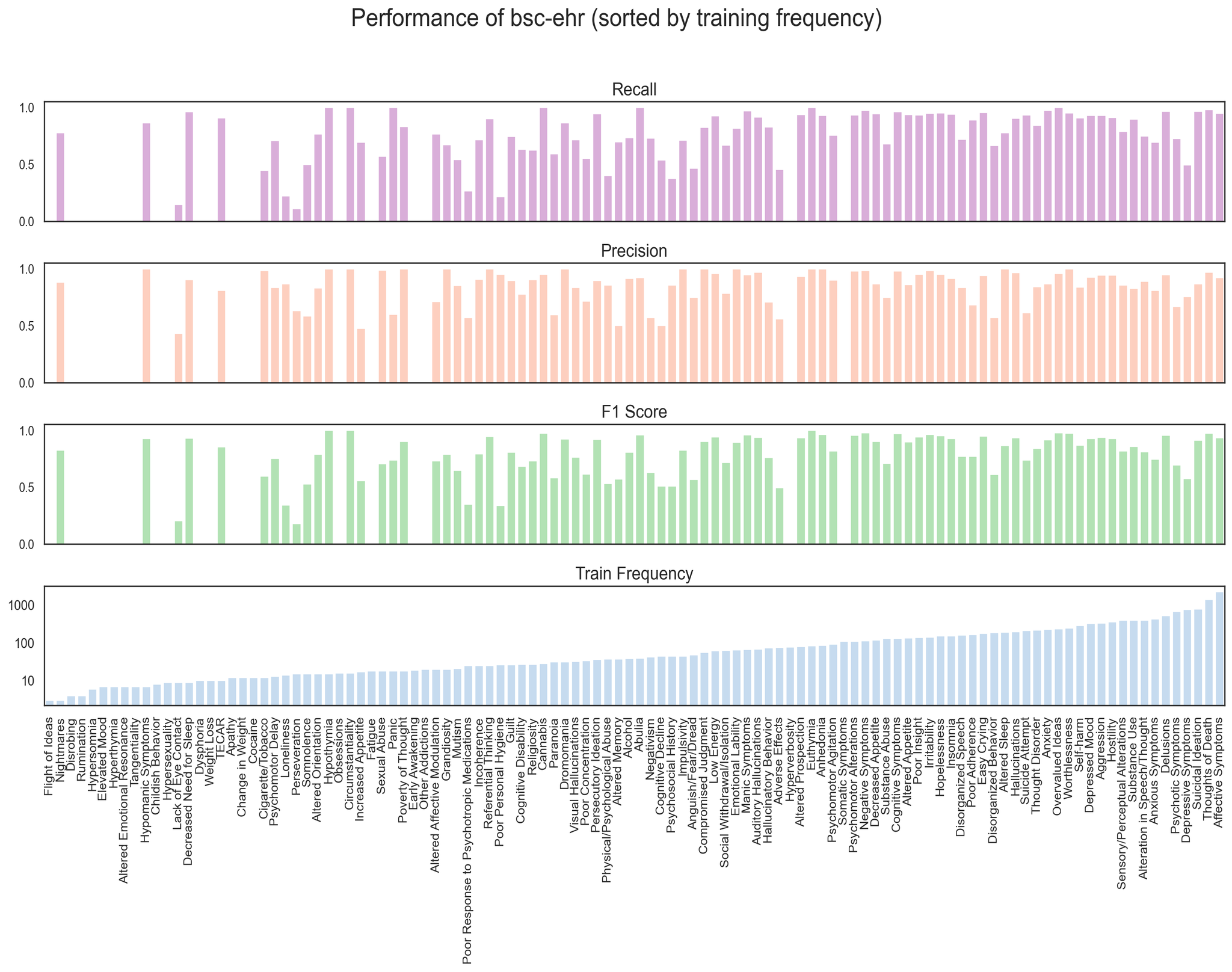

**Supplementary Figure 3**. Per-concept performance of bsc-ehr on the CSJDM training set, evaluated on the CSJDM test set (n=358 documents). The three performance panels show per-concept recall (top, pink), precision (middle, orange), and F1 score (bottom, green). The bottom panel shows the training set frequency for each concept on a log_10_ scale (blue). Concepts are ordered by their training frequency (increasing left to right).

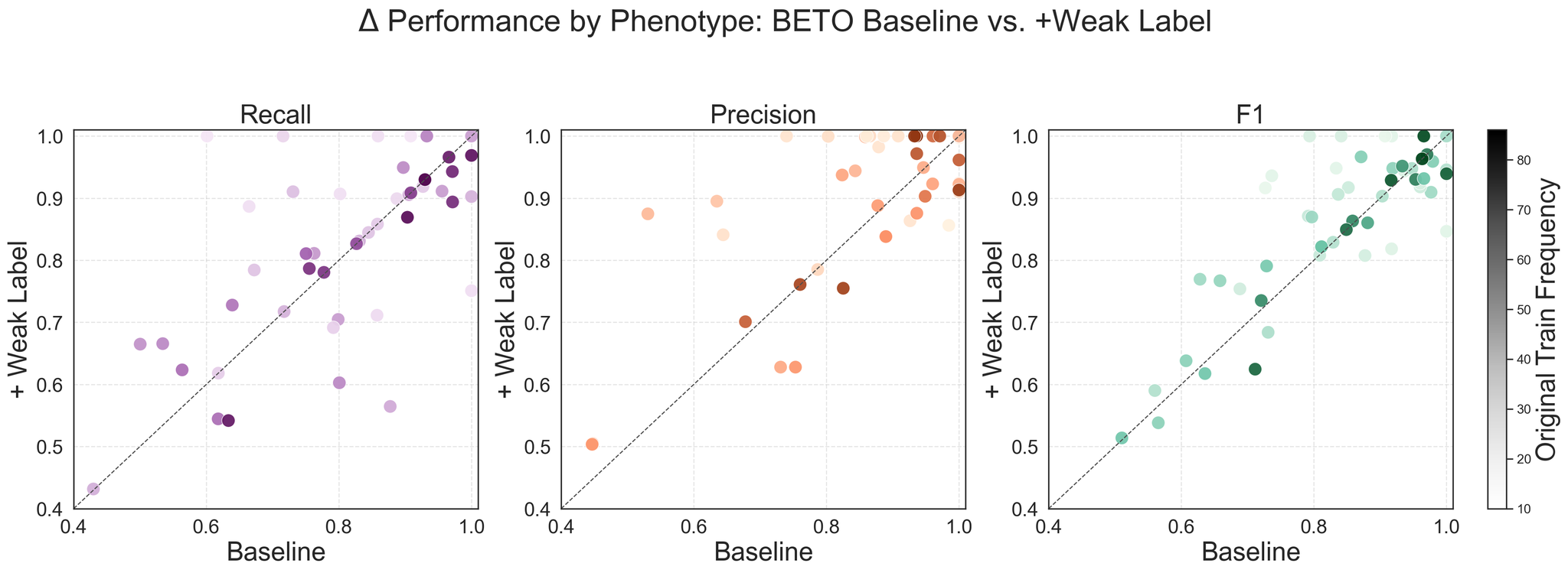

**Supplementary Figure 4.** Comparison of BETO-Baseline and BETO +WL concept-level performance across concepts. Each panel displays the relationship between baseline and weak-label models for recall (left, purple), precision (middle. orange), and F1 (right, green). The color intensity encodes the original training frequency of that concept (lighter = fewer training examples; darker = more training examples). Points above the dashed identity line (y=x) represent concepts that improved with weak-label augmentation. Analysis includes concepts with less than 100 training set frequency in the CSJDM training set, and had a tNLP algorithm developed (N concepts = 55).

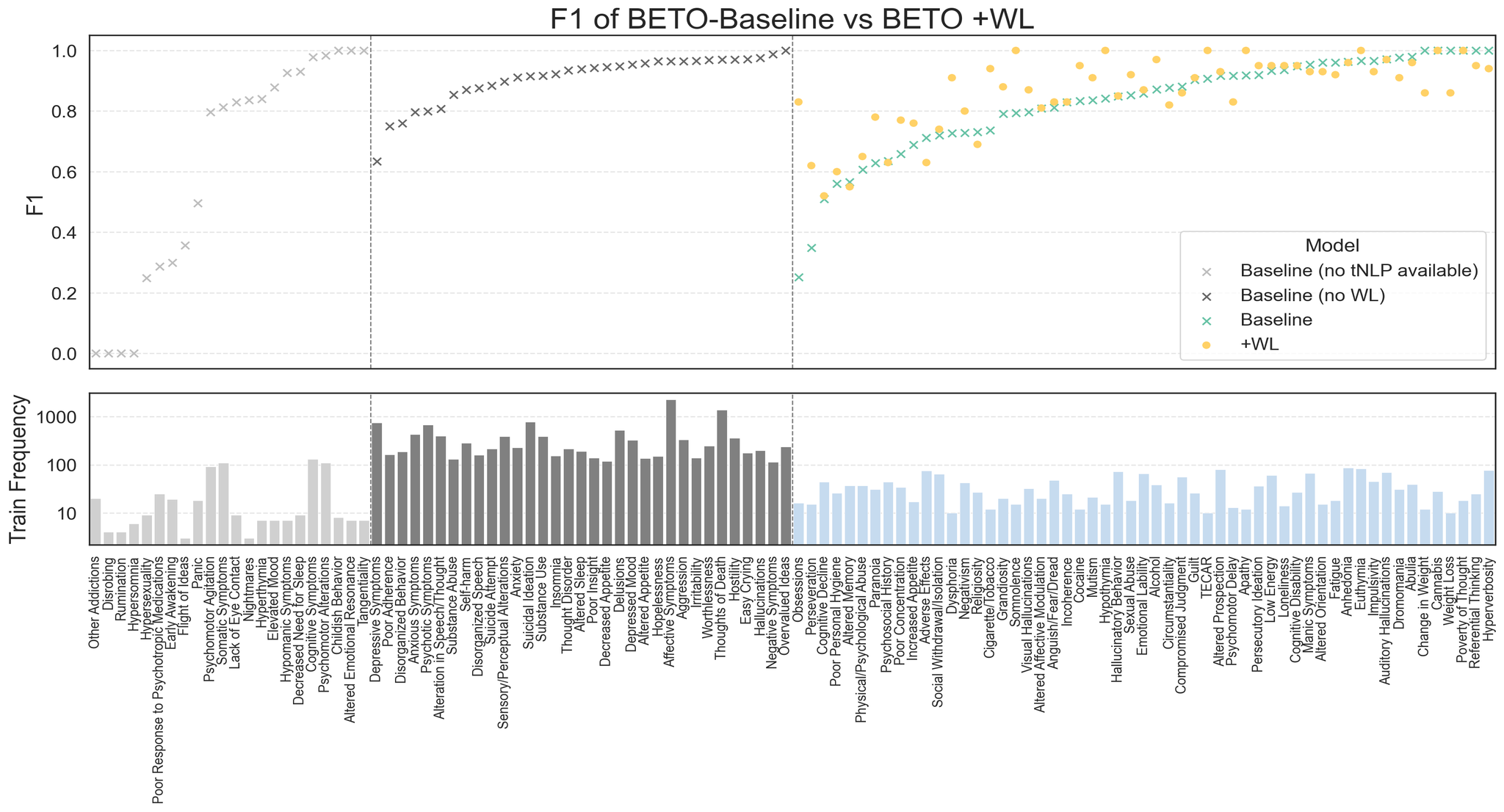

**Supplementary Figure 5**. Concept-level performance of BETO-Baseline vs. BETO +WL across concepts. The top panel shows F1 scores, where crosses represent BETO-Baseline’s performance and yellow circles represent BETO +WL. Concepts are grouped into three regions: (left) concepts without a tNLP method, (middle) concepts with more than 100 training set examples in the CSJDM dataset, and (right) concepts fine-tuned with weak labels. The bottom panel shows the training set frequency on a log_10_ scale for the corresponding concepts. Concepts are ordered by their training frequency (increasing left to right).

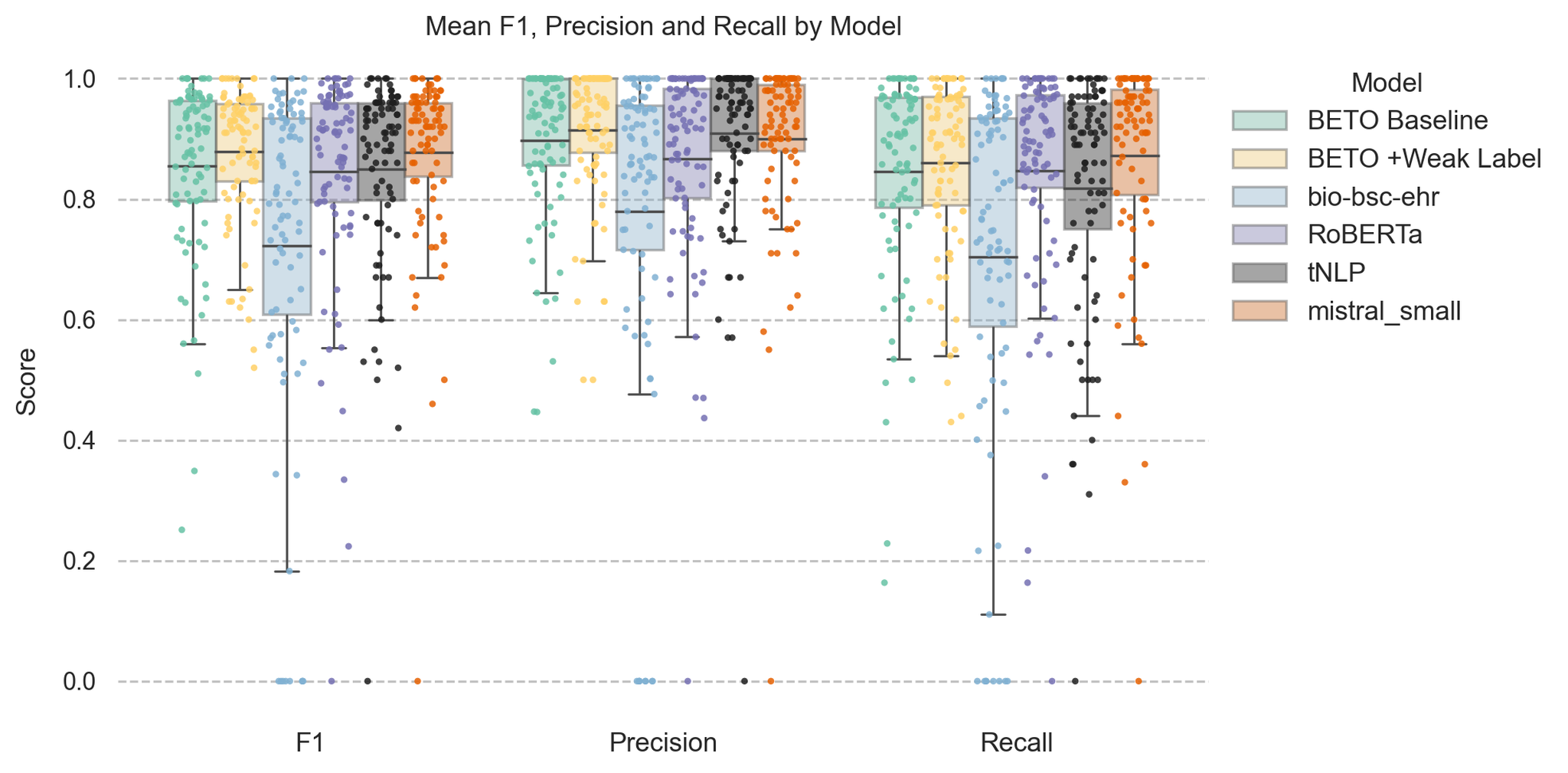

**Supplementary Figure 6**. Distribution of concept-level F1, precision, and recall scores across six models. Boxplots and overlaid scatter points show performance variability across concepts for BETO-Baseline (green), BETO +WL (yellow), bsc-ehr (blue), XLM-RL (purple), tNLP patterns (black), and fine-tuned Mistral-small (orange). Boxplots show the interquartile range (IQR) with whiskers extending to 1.5×IQR, and horizontal gray ticks mark the mean score. All model performances were reported on the set of 88 concepts for which the tNLP method was evaluated.

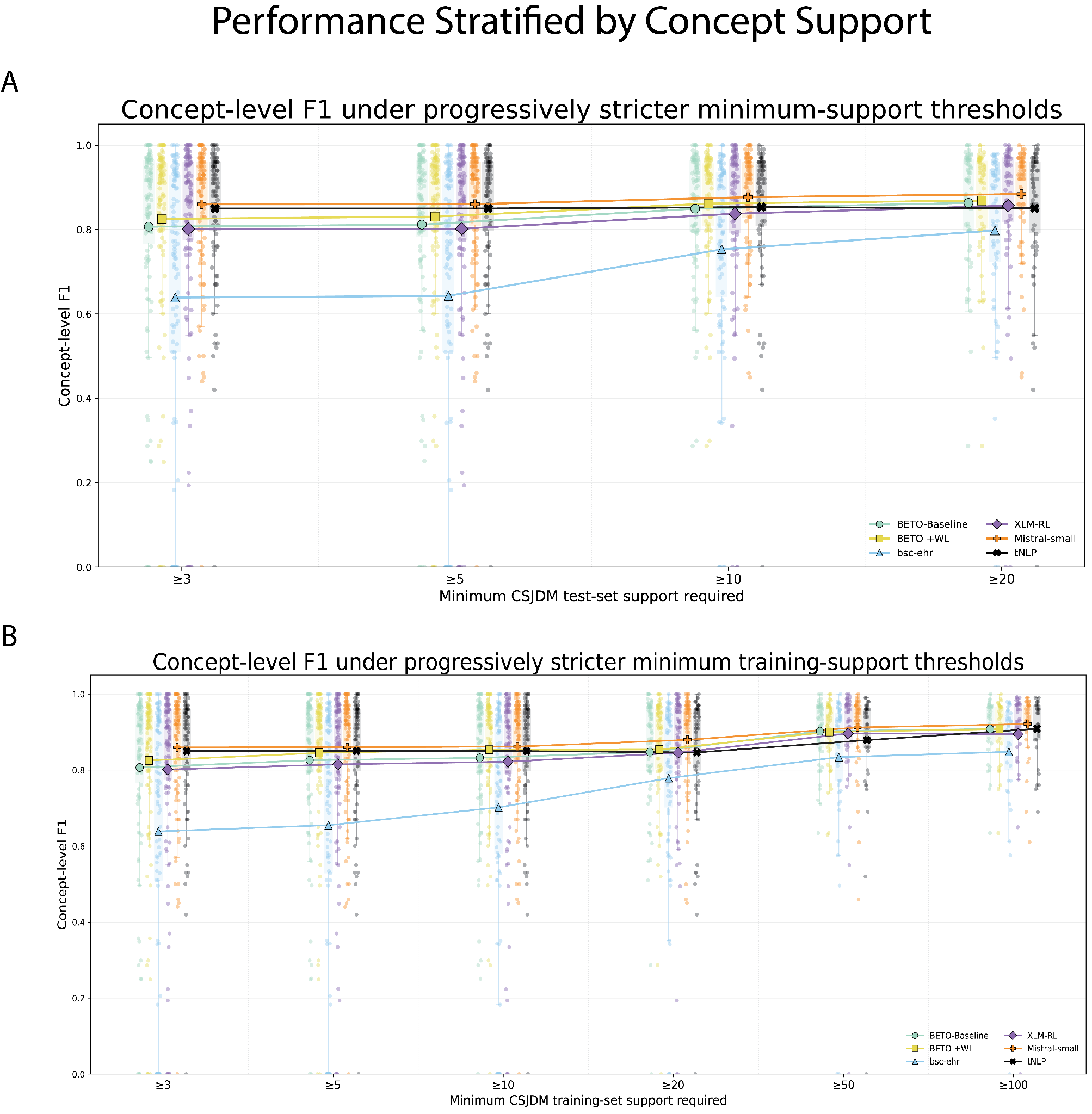

**Supplementary Figure 7. Concept-level F1 performance across progressively stricter support thresholds.**

A) Concept-level F1 distributions after restricting evaluation to concepts with at least 3, 5, 10, or 20 instances in the CSJDM test set.

(B) Concept-level F1 distributions after restricting evaluation to concepts with at least 3, 5, 10, 20, 50, or 100 positive mentions in the CSJDM training set.

Grouped dotplots show performances for BETO-Baseline (green), BETO +WL (yellow), bsc-ehr (blue), XLM-RL (purple), tNLP patterns (black), and fine-tuned Mistral-small (orange). Faint points represent individual concept-level F1 scores, faint boxes indicate the interquartile range, filled markers indicate the mean F1 for each model within that support threshold, and lines connect model-specific means across thresholds. Thresholds are cumulative, such that concepts meeting a higher threshold are also included at all lower thresholds. At each threshold, calculations include only concepts for which performance estimates were available for the corresponding model.

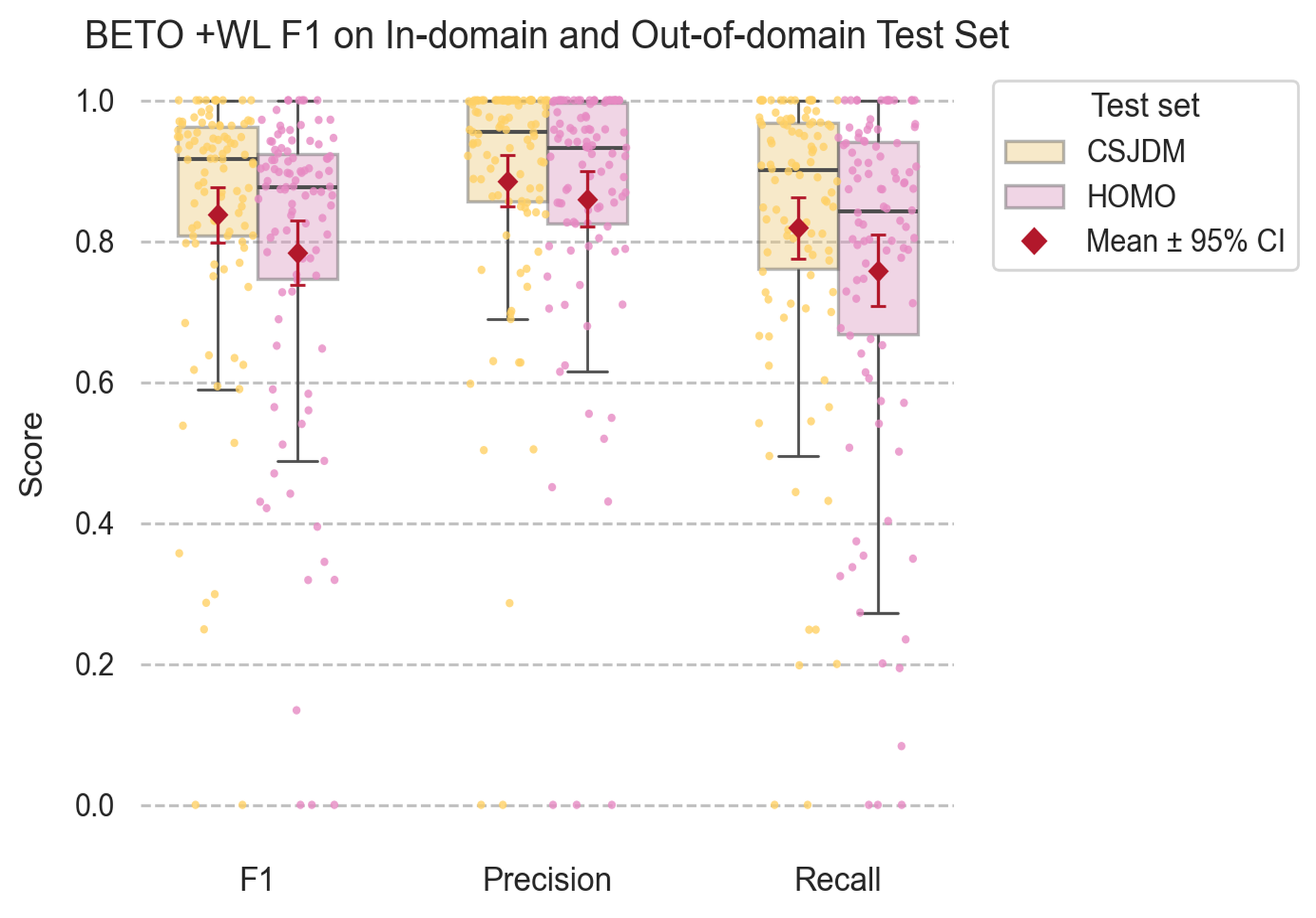

**Supplementary Figure 8.** Distribution of concept-level F1, precision, and recall scores for BETO +WL on the CSJDM (yellow) and HOMO (pink) test sets. Each point represents performance for a single concept (total concepts = 98). Boxplots show the median and interquartile range (IQR) across concepts, with whiskers extending to 1.5×IQR. Red diamonds and error bars indicate the mean and 95% confidence interval.

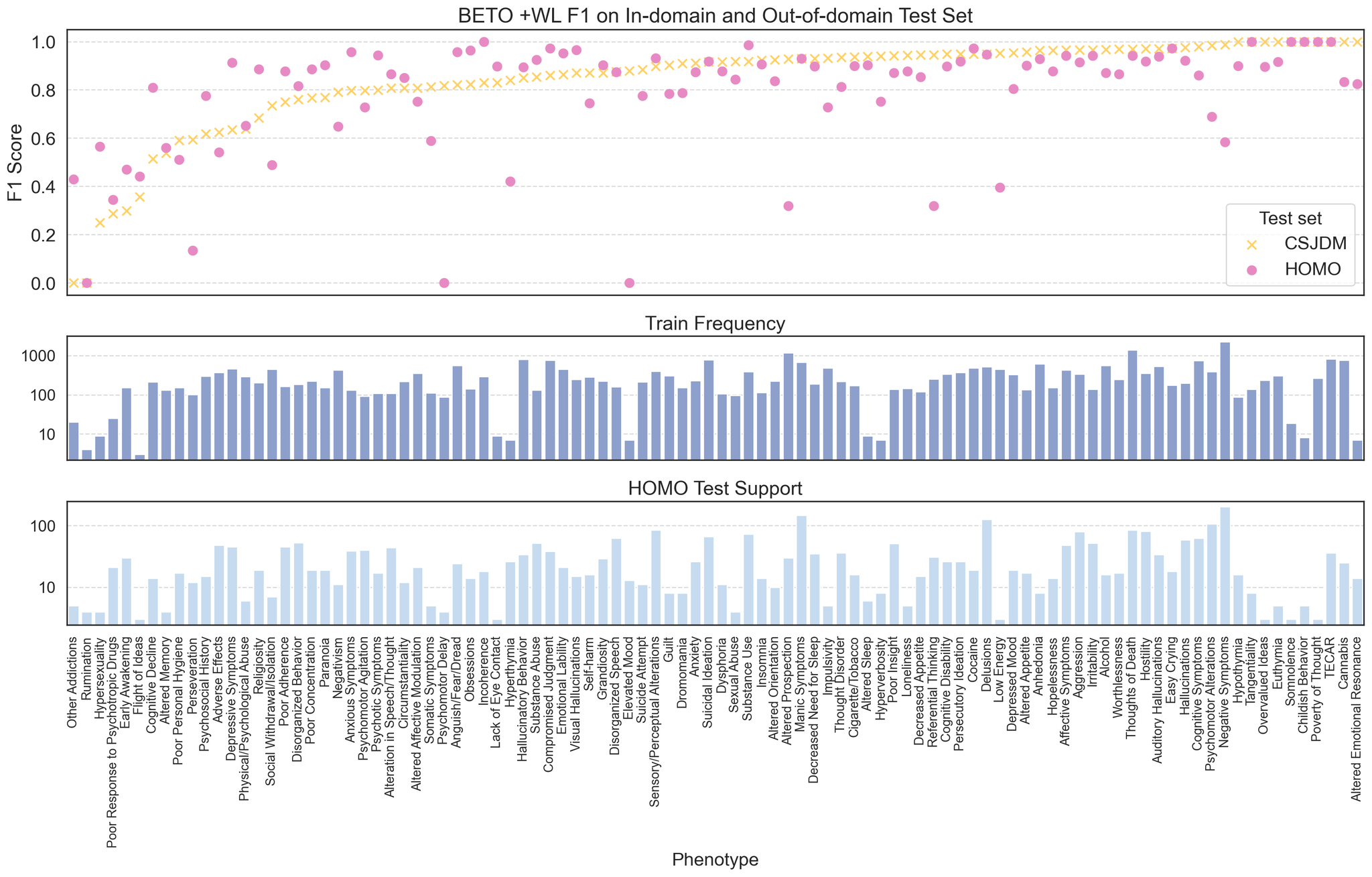

**Supplementary Figure 9**. Cross-testing performance of BETO +WL on CSJDM and HOMO test sets by concept. Top panel: F1 scores for BETO evaluated on the CSJDM test set (yellow crosses) and the HOMO test set (pink circles). Only concepts with at least three test examples in HOMO were included in the evaluation. Middle panel (dark blue): training set frequency on a log₁₀ scale for each concept in the CSJDM dataset. Bottom panel (light blue): test set support on a log₁₀ scale for concepts in the HOMO dataset. Concepts are shown in ascending order of BETO +WL F1 score on CSJDM. Only concepts with at least three instances in the HOMO test set are included (N concepts = 98).

**Supplementary Notes**

**Supplementary Note 1. Weight Assignment**

| tNLP Precision | Strong Labels Weight | Weak Labels Weight |
| --- | --- | --- |
| Precision < 0.75 | 5 | 0.2 |
| 0.75 <= Precision < 0.9 | 5 | 0.5 |
| Precision >= 0.9 | 5 | 1 |

**Supplementary Note 2. Model Selection**

BETO

BETO is a BERT-base model with 110M parameters, pretrained on the Spanish Unannotated Corpora (SUC), which consists of approximately 3 billion words (~18 GB) of raw Spanish text. Despite its relatively compact size, BETO has consistently shown performance comparable to that of much larger multilingual models, while also outperforming certain domain-specific Spanish models, making it a strong general-purpose baseline for this study [34].

bsc-bio-ehr-es

The bsc-bio-ehr-es (hereafter: “bsc-ehr”) model is a RoBERTa-based encoder with 125M parameters, pretrained on a large-scale biomedical and clinical corpus in Spanish comprising more than 278,000 clinical documents. Evaluations have shown that it outperforms both general-domain and Spanish clinical BERT variants on several clinical named entity recognition (NER) tasks, achieving higher F1 scores [32].

XLM-RoBERTa-large

XLM-RoBERTa-large (hereafter: “XLM-RL”) is a multilingual RoBERTa model with 550M parameters, pretrained on 2.5 TB of filtered CommonCrawl data spanning 100 languages. Approximately 2% of this corpus, corresponding to ~53 GB of text, consisted of Spanish. Released by Facebook AI in 2020, the model demonstrated performance comparable to monolingual BERT variants across a wide range of downstream benchmarks, underscoring its strong cross-lingual transfer capabilities.

**Supplementary Note 3. Fine-Tuning with Annotated Data**

Fine-tuning example for the concept *Alcohol*

| Token Sequence | ['[CLS]', 'paciente', 'de', '3', '##6', 'años', 'de', 'edad', ',', 'con', 'antecedentes', 'descritos', ',', 'ya', 'conocido', 'en', 'la', 'institu', '##cion', ',', 'ingresa', 'bajo', 'efecto', 'de', 'consumo', 'de', 'spa', 'y', **'alcohol'**, ',', 'durante', 'entrevista', 'con', 'llanto', ',', 'verbal', '##iza', 'ideas', 'de', 'muerte', 'e', 'idea', '##cion', 'suicida', ',', 'manifiesta', 'deseo', 'de', 'iniciar', 'proceso', 'de', 'desin', '##tox', '##ica', '##cion', ',', 'por', 'lo', 'cual', 'en', 'el', 'momento', 'se', 'indica', 'hospital', '##izar', '.', '[SEP]', '[PAD]', '[PAD]', ......, '[PAD]'] |
| --- | --- |
| Annotations | [ O, O, O, O, …, **I-Alcohol**, O, O, …, O] |
| Labels | [-100, 0, 0, 0, 0, ……, **1**, 0, 0, 0, ……., -100, -100, -100] |

Fine-tuning hyperparameters

All models were fine-tuned for 10 epochs using the hyperparameters summarized in the table below. The configuration was selected to balance sufficient training with the risk of overfitting, given the absence of a development set for early stopping. Training was conducted with a batch size of 16, consistent with recommendations from the original BERT publication and well-suited to the limited size of the dataset. The learning rate was fixed at 1e-5, a standard value for transformer fine-tuning that provides stable convergence under conditions of sparse positive labels. Maximum input length was set to 512 tokens, the default for BERT, in order to minimize truncation of long psychiatric sentences and multi-clause spans. All other hyperparameters followed the default configurations provided by the Hugging Face Transformers library.

| **Hyperparameter** | **Value** |
| --- | --- |
| Number of Epochs | 10 |
| Batch Size | 16 |
| Maximum sequence length | 512 tokens |
| Optimizer | AdamW |
| Learning Rate | 1×10⁻⁵ |
| Warmup steps | 0 |
| Gradient Accumulation Steps | 1 |
| Weight Decay | 0.0 |
| Adam β₁ | 0.9 |
| Adam β₂ | 0.999 |
| Adam ε (epsilon) | 1×10⁻⁸ |
| Maximum Gradient Norm | 1.0 |
| Early Stopping | Not used |

**Supplementary Note 4. Fine-Tuning with Weak Labels**

Weight Integration into the Loss Function

We implemented per-example weighting by modifying the Hugging Face *Trainer*. During training, the model first computes token-level cross-entropy losses for every position in the sequence. Losses from padding or special tokens are masked out, and the remaining token losses are averaged within each sentence to obtain a single loss value per example.

Each example is then scaled by its assigned weight, which reflects annotation quality: clinician-annotated sentences always receive a higher fixed weight, while weakly labeled sentences are weighted according to the precision of the pattern-matching algorithm used to generate their labels. Since all tokens in a sentence share the same source, they are assigned the same weight. Finally, the weighted example losses are averaged across a batch of 16, ensuring that high-quality annotations have stronger influence on model optimization while still allowing precise weak labels to contribute signal.

Example of Per-Example/Sentence Weighted Loss Computation

| **Input Annotation Type** | Weak label |
| --- | --- |
| **Pattern-Match Precision** | 0.78 |
| **Token Sequence** | ['[CLS]', 'actualmente', 'no', 'esta', 'incapa', '##cita', '##do', 'asiste', 'al', 'trabajo', 'para', 'cumplimiento', 'del', 'horario', ',', 'no', 'desempeña', 'ninguna', 'función', ',', 'situación', 'que', 'refuerza', 'sus', 'cogni', '##ciones', 'de', 'inutil', '##idad', '.', '[SEP]', '[PAD]', ..., '[PAD]'] |
| **Raw Per-Token Loss** | [0.0000, 0.8514, 0.8600, 0.6673, 0.6034, 0.6627, 0.8642, 0.6562, 0.6738, 1.1703, 0.8196, 1.0252, 0.8347, 0.5361, 0.4167, 0.6006, 0.8343, 0.7779, 0.6534, 0.2934, 0.4138, 0.3901, 0.5248, 0.4259, 0.3321, 0.4485, 0.3597, 0.6765, 0.6088, 0.5556, 0.0000,..., 0.0000] |
| **Raw Per-Sentence/**  **Example Loss** | 0.6392 |
| **Per-Sentence/**  **Example Weight** | 0.5 |
| **Weighted Per-Sentence/**  **Example Loss** | 0.3196 |
